# Variability in mRNA SARS-CoV-2 BNT162b2 vaccine immunogenicity is associated with differences in the gut microbiome and habitual dietary fibre intake

**DOI:** 10.1101/2022.08.24.22279143

**Authors:** Genelle R. Healey, Liam Golding, Alana Schick, Abdelilah Majdoubi, Pascal M. Lavoie, Bruce A. Vallance

## Abstract

**Objective:** Little is known about the interplay between gut microbiome and SARS-CoV-2 vaccine immunogenicity. In this prospective observational study, we investigated associations between the gut microbiome, habitual dietary fibre intake, and mRNA vaccine-elicited immune responses, including anti-Spike IgG, avidity, and ACE-2 competition (surrogate neutralization).

**Design:** 16S rRNA sequencing and short-chain fatty acid analyses were undertaken using stool samples collected from 48 healthy individuals at baseline and twelve-weeks after 1^st^ BNT162b2 SARS-CoV-2 vaccine dose. Associations between gut microbiome data and SARS-CoV-2 spike and RBD IgG levels, competitive binding antibodies, and anti-SARS-CoV-2 spike total relative fractional avidity assays were evaluated. A validated dietary fibre intake food frequency questionnaire was also used to correlate habitual dietary fibre intakes with vaccine responses.

**Results:** Our data revealed several baseline bacterial taxa, including *Prevotella, Haemophilus* and *Veillonella* (p<0.01), associated with BNT162b2 vaccine responses. Several *Bacteroides* spp. (p<0.01) as well as *Bifidobacterium animalis*, (p=0.003), amongst others, were positively associated with antibody avidity. Conversely, concentrations of isovaleric and isobutyric acid were higher in individuals with the lowest SARS-CoV-2 vaccine responses (p<0.01). Classifying participants based on habitual dietary fibre intake identified distinct avidity responses.

**Conclusion:** We showed associations between baseline gut microbiota composition and immunogenicity of BNT162b2 vaccine responses, particularly avidity maturation. We also demonstrate that branched-chain fatty acids and habitual dietary fibre intakes are associated with BNT162b2 vaccine immunogenicity. Together these findings indicate a link between gut microbiome, diet and antibody immunity to SARS-CoV-2 spike protein, suggesting interventions which modulate the gut microbiome could enhance COVID-19 vaccine responses.

**SIGNIFICANCE OF THIS STUDY:** *What is already known on this subject?:*

- Strength and persistence of the SARS-CoV-2 BNT162b2 vaccine is variable between individuals.
- To date, only one study has demonstrated that baseline gut microbiota can predict SARS-CoV-2 vaccine response.

*What are the new findings?:*

- For the first time we showed that the higher concentrations of branched-chain fatty acids, isovaleric and isobutyric acids, are negatively associated with SARS-CoV-2 BNT162b2 vaccine responses.
- We revealed that habitual dietary fibre intake led to variability in the strength of antibody binding after the BNT162b2 vaccine. Specifically, high dietary fibre consumers displayed a significant increase in antibody avidity between in their 1^st^ and 2^nd^ dose.

*How this study might affect research, practice, or policy:*

- Our data suggests that therapeutic interventions which target the gut microbiome, including dietary modification, as well as pre-, pro-, and post-biotics, could enhance BNT162b2 vaccine immunogenicity, thus helping in the fight against COVID-19.

## INTRODUCTION

A key strategy for controlling the COVID-19 pandemic is conferring robust and sustained immunity via the administration of SARS-CoV-2 vaccines. Novel strategies to control the spread of the SARS-CoV-2 virus and boost the immunogenicity of vaccines will prove helpful in the fight against COVID-19. mRNA vaccines offer high efficacy in protecting against severe disease caused by the wild-type strain and Omicron subvariants^1^. However, antibody responses to FDA-approved SARS-CoV-2 vaccines are extremely heterogeneous with some individuals failing to mount high quality responses^2,3^. Moreover, the vaccines do not provide sustained protection against re-infection or symptomatic infection^4^. Therefore, it is important to understand the factors influencing the immunogenicity of SARS-CoV-2 vaccines, including their capacity for long-term protection.

Factors such as age, chronic disease, smoking, body mass index, depression, and stress have all been implicated in vaccine responses, including those targeting SARS-CoV-2^5–8^. Interestingly, the factors outlined above have also been shown to impact the gut microbiome. It is, therefore, plausible that differences in gut microbiome profiles between individuals could contribute to inter-individual variability in observed vaccine responses. Elucidating the role the gut microbiome plays in SARS-CoV-2 vaccine immunogenicity offers the potential to screen for potentially poor vaccine responders, as well as utilizing microbiome-targeted therapies, i.e., prebiotics, probiotics and postbiotics^9^, as adjuvants to enhance vaccine responses.

To date, only one publication has investigated potential associations between the gut microbiome and SARS-CoV-2 vaccine immunogenicity^8^. Ng and colleagues associated *Bifidobacterium adolescentis* with higher antibody responses after whole inactivated SARS-CoV-2 CoronaVac vaccination. This study also found a positive association between neutralising antibodies elicited by BNT162b2 vaccination and the presence of bacteria that express adhesive surface structures such as flagella and fimbriae. While informative, additional research is clearly needed to further elucidate the role that specific gut microbiota and associated metabolites (i.e., short-chain fatty acids [SCFA]) play in the immunogenicity of SARS-CoV-2 vaccines.

In this study, we investigated the impact of habitual dietary fibre intakes, gut microbiome signatures, and SCFAs, on mRNA (Pfizer-BioNTech BNT162b2) vaccine responses after the 1^st^ and 2^nd^ doses.

## MATERIALS AND METHODS

### Study cohort

Uninfected healthcare workers aged ≥ 18 years were prospectively recruited at British Columbia Children’s Hospital (Vancouver, Canada) between February and August 2021 prior to receiving two Pfizer-BioNtech BNT162b2 mRNA vaccine doses, approximately 4 months apart. Participants who tested positive for SARS-CoV-2 by viral or Spike antibody testing of baseline samples or nucleocapsid antibody testing of any sample were excluded. Blood samples were collected at baseline (pre-vaccine), 12 weeks after 1^st^ vaccination, and 4 weeks after 2^nd^ Pfizer-BioNtech BNT162b2 mRNA vaccination. Stool samples were collected at baseline and 12 weeks after the 1^st^ vaccine dose (**figure 1A**). This study was approved by the University of British Columbia Clinical Research Ethics Board (H20-01205). Informed e-consent for collection of blood and stool samples was obtained from all participants. Information on the participant characteristic data collected can be found in the online supplementary information.

**Figure 1.**
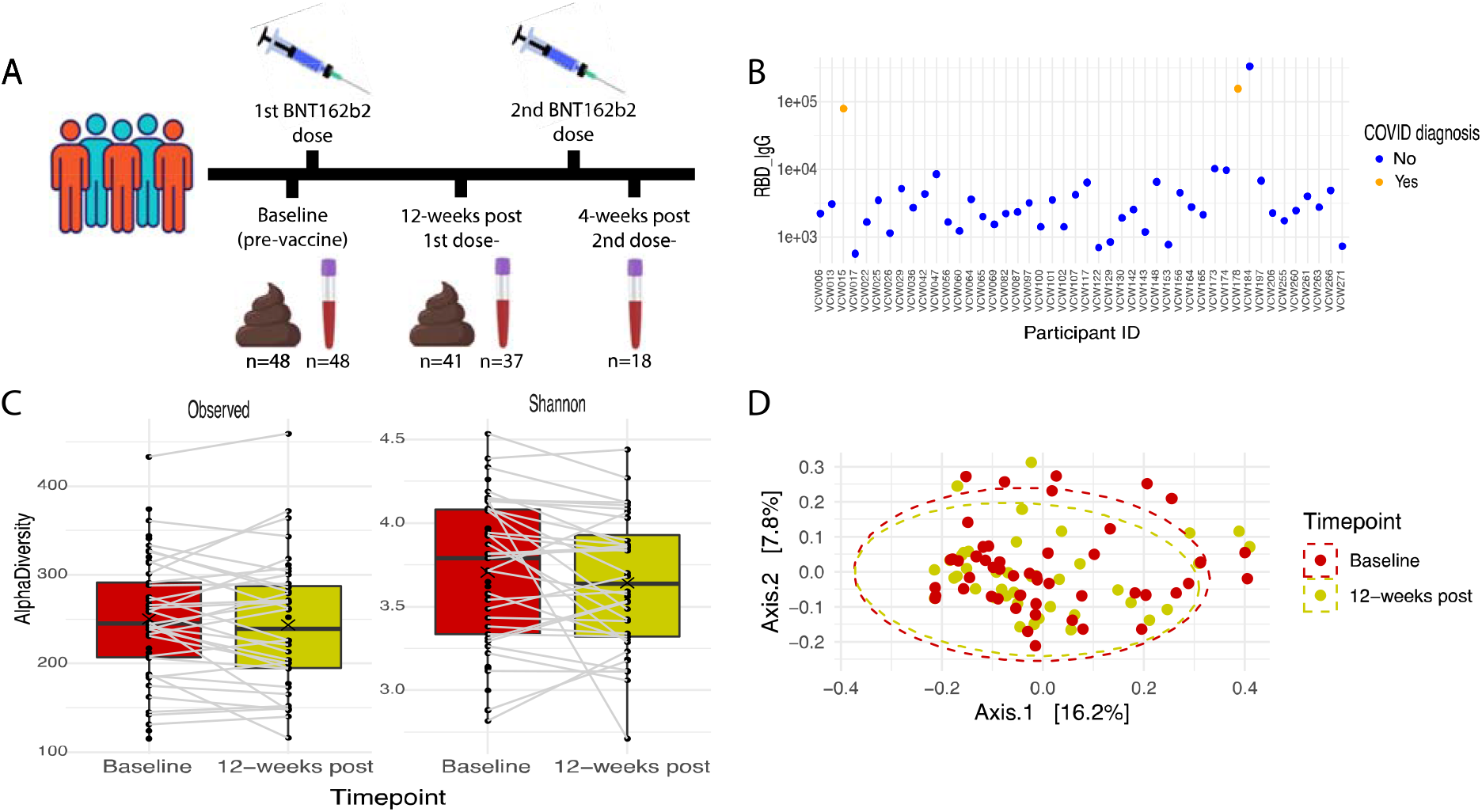
Participant flow through the study, COVID-19 diagnosis and changes in alpha and beta diversity from baseline to twelve-weeks post 1^st^ BTN162b2 vaccine. (A) Study design and sample collection. (B) Three participants were excluded from the analyses as two participants indicated a positive SARS-CoV-2 test during the study and one participant may have had a missed COVID-19 diagnosis due to a high RBD IgG level. (C) A trend towards a significant reduction in Shannon alpha diversity was observed (p=0.07812; Wilcoxon rank test). (D) A significant shift in beta diversity occurred between baseline and post 1st BTN162b2 vaccine (p=0.0159; PERMANOVA).

### Serum sample analyses

#### SARS-CoV-2 spike and RBD IgG multiplex assay

Anti-SARS-CoV-2 spike and receptor-binding domain (RBD)-specific antibodies were quantified using a multiplexed electro-chemiluminescent assay (MSD, Rockvile, MD)^10^. Details relating to this assay can be found in the online supplementary information.

#### SARS-CoV-2 spike and RBD ACE-2 competition assay

ACE-2 competitive binding (U/ml) was quantified using the same MSD multiplex assay as described above. A detailed description of the method can be found in the online supplementary information.

#### Anti-SARS-CoV-2 spike absolute and relative fractional avidity assay

Anti-spike absolute and relative fractional avidities were determined using a modified version of a previously described method^11,12^. The full methodology can be found in the online supplementary information.

### Stool collection

Participants were provided with a stool sample collection kit and written instructions at their initial and twelve-week post 1^st^ BNT162b2 vaccine clinic visits. Details relating to how stool samples were collected can be found in the online supplementary information.

### Short-chain fatty acid analysis

Stool samples, collected in DNA Genotek ME-200 tubes (80-95% ethanol), were thawed on ice, vortexed, and 700 μl was transferred to a Savant SPD131 DDA speedvac (Thermo Scientific) centrifugal vacuum concentrator and run at room temperature until virtually all the ethanol had evaporated. The samples were then processed as previously described in the online supplementary information.

### 16S rRNA sequencing

The ThermoFisher MagMax microbiome nucleic acid ultra-isolation kit and semi-automated Kingfisher Duo Prime were used to extract microbial DNA from the stool samples following the manufacturer’s instructions with minor modifications. 16S library preparation for individual samples were performed at the BC Children’s Hospital Research Institute Gut4Health Microbiome Sequencing CORE, using a similar method to Kozich and colleagues^13^. Further details relating to 16s rRNA sequencing can be found in the online supplementary information.

### Statistical analyses

Refer to the online supplementary information for details relating to the statistical analyses undertaken.

## RESULTS

### Study cohort

In total, 48 participants had a baseline stool sample collected and 41 had a sample collected at twelve-weeks post 1^st^ Pfizer-BioNTech BTN162b2 mRNA vaccine. Of these participants, 38 had matching stool samples, and 37 and 18 had blood samples collected twelves-weeks post 1^st^ and 4 weeks post 2^nd^ BTN162b2 vaccine doses, respectively (**table S1, figure 1A**). The clinical characteristics of the 48 participants are shown in **table S1**. Two additional participants (beyond the 48) tested positive for SARS-CoV-2 during the study and were excluded from subsequent analyses. One additional participant was also excluded as a possible SARS-CoV-2 misdiagnosis indicated by a high RBD IgG level (**figure 1B**).

### Gut microbiome changes post 1^st^ dose of BNT162b2 vaccine

We undertook 16S rRNA sequencing on the 48 baseline and 41 twelve-week post 1^st^ BTN162b2 vaccine stool samples. We generated on average 24.67 Mb (90,509 reads) of data per sample. We also submitted 42 baseline and 37 twelve-week post 1^st^ BTN162b2 vaccine stool samples for SCFA analysis. Participant-specific antibiotic use as well as changes in weight and diet were reported as these are common confounding factors in gut microbiota analyses (**table S1**). Only a small number of participants reported taking antibiotics (n=3) or experiencing changes in weight or diet (n=7). As the data from these participants were not considered outliers they were included in subsequent analyses.

First, we compared the changes in gut microbiota alpha and beta diversity from baseline to twelve weeks post 1^st^ vaccine. There was a trend towards a reduction in Shannon alpha diversity (p=0.07812; Wilcoxon) after the 1^st^ vaccine dose (**figure 1C**). As well, a significant shift (p=0.0159; PERMANOVA) in the gut microbiota community (i.e., beta diversity) was observed from baseline to after the 1^st^ dose (**figure 1D, figure S1A-C**). The specific microbes that were changed after the 1^st^ dose included a significant reduction in Actinobacteriota (p<0.0001, log_2_(FoldChange) = -0.672; Wald test), *Anaerostipes* (p=0.00161, log_2_(FoldChange) = -0.745; Wald test) and *Blautia* (p=0.00103, log_2_(FoldChange) = -0.548; Wald test) and an increase in *Lachnoclostridium* (p=0.0018, log_2_(FoldChange) = -0.448; Wald test) (**figure 2A**). A previous study undertaken in a cohort in Hong Kong^8^ also demonstrated that the BNT162b2 vaccine led to a significant reduction in Actinobacteriota and *Blautia* spp. Although significant changes in several bacterial taxa were observed, no significant changes (p>0.05; Wilcoxon) in SCFAs were shown after the 1^st^ vaccine dose (**figure 2B**).

**Figure 2.**
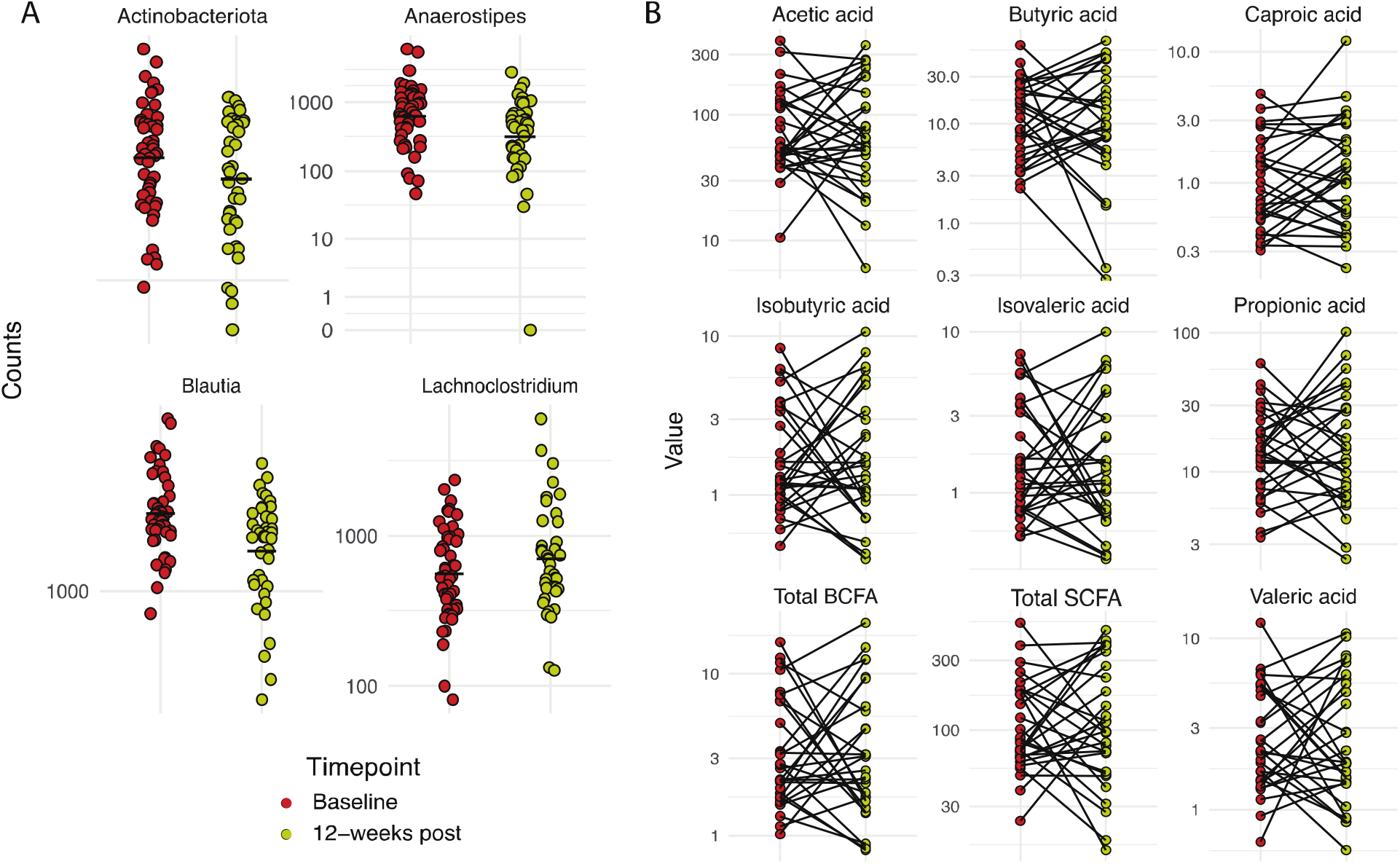
Change in specific taxonomy and SCFA production from baseline to twelve-weeks post 1^st^ BTN162b2 vaccine. (A) A significant reduction in Actinobacteriota (p<0.0001), *Anaerostipes* (p=0.00161) and *Blautia* (p=0.00103) an increase in *Lachnoclostridium* (p=0.00179) were observed after the 1^st^ BNT162b2 vaccine. (B) No significant changes (p>0.05) in SCFAs were observed after the 1^st^ BNT162b2 vaccine. P-values determined using Wald test of significance.

### Baseline gut microbiota and SCFAs impact immune response to BNT162b2 vaccination

Ng and colleagues^8^ showed that specific baseline gut microbiota taxa impact immune responses to the BNT162b2 vaccine. Therefore, we assessed each participants response to the 1^st^ BTN162b2 vaccine dose using relevant immune parameters including anti-RBD and spike IgG levels, ACE-2 competitive binding antibody levels, and the anti-RBD and spike IgG to competitive binding antibody ratio (measure of neutralizing capacity per antibody). Following SARS-CoV-2 Spike mRNA vaccination many individuals develop antigen-specific B cells which undergo class-switching and somatic hypermutation to produce IgG antibodies^14^. To assess the relationship of the microbiome with immune outcomes, we selected parameters previously shown to correlate with effective vaccine responses. Anti-Spike and anti-RBD IgG levels regulate the magnitude of *in vitro* antibody-dependent functions such as complement deposition, Fc-mediated cellular cytotoxicity and phagocytosis as well as live virus neutralization^15–17^. To specifically measure the capacity to block RBD-ACE-2 binding, we performed a commercially available *in vitro* anti-Spike and -RBD surrogate neutralization ACE-2 competition assay which has been shown to strongly correlate with live virus neutralization^18^. To evaluate the potency or the capacity of anti-Spike/RBD IgG to mediate ACE-2 blocking, we calculated a ratio of ACE-2 competitive binding antibodies by IgG for both Spike and RBD.

Variability in RBD IgG to competitive binding antibody ratio between participants was the immune parameter most strongly associated (p=0.07; Wald test) with variation in the gut microbiota (**figure 3A, figure S2A**). There were, however, specific taxa that were significantly associated (p<0.05; Wald test) with several vaccine response parameters (**figure 3B, figure S2B-C**). The most pronounced associations included a positive correlation between twelve-week post 1^st^ BNT162b2 vaccine RBD IgG levels and baseline *Prevotella* counts (p=0.032; Wald test) (**figure 3C**) and negative correlations between post 1^st^ vaccine RBD competitive binding antibody levels and baseline *Clostridiales* bacterium DTU089 and CAG-352, and *Megasphaera* counts (p=0.038, p=0.038 and p=0.047, respectively; Wald test) (**figure 3D, figure S2B**). Counts of *Clostridiales* bacterium DTU089 at baseline were also negatively correlated with post 1^st^ vaccine spike competitive binding antibody levels (p=0.029; Wald test) (**figure 3E, figure S2C**). Lastly, baseline Shannon and observed alpha diversity metrics were negatively correlated with spike IgG (p=0.04, r= -0.3; Spearman rank) (**figure S3A**), and RBD competitive binding antibody (p=0.035, r= -0.31; Spearman rank), RBD IgG (p=0.04, r= -0.3; Spearman rank), spike competitive binding antibody (p=0.0077, r= -0.4; Spearman rank) and spike IgG (p=0.0073, r= -0.4; Spearman rank), respectively (**figure S3B**).

**Figure 3.**
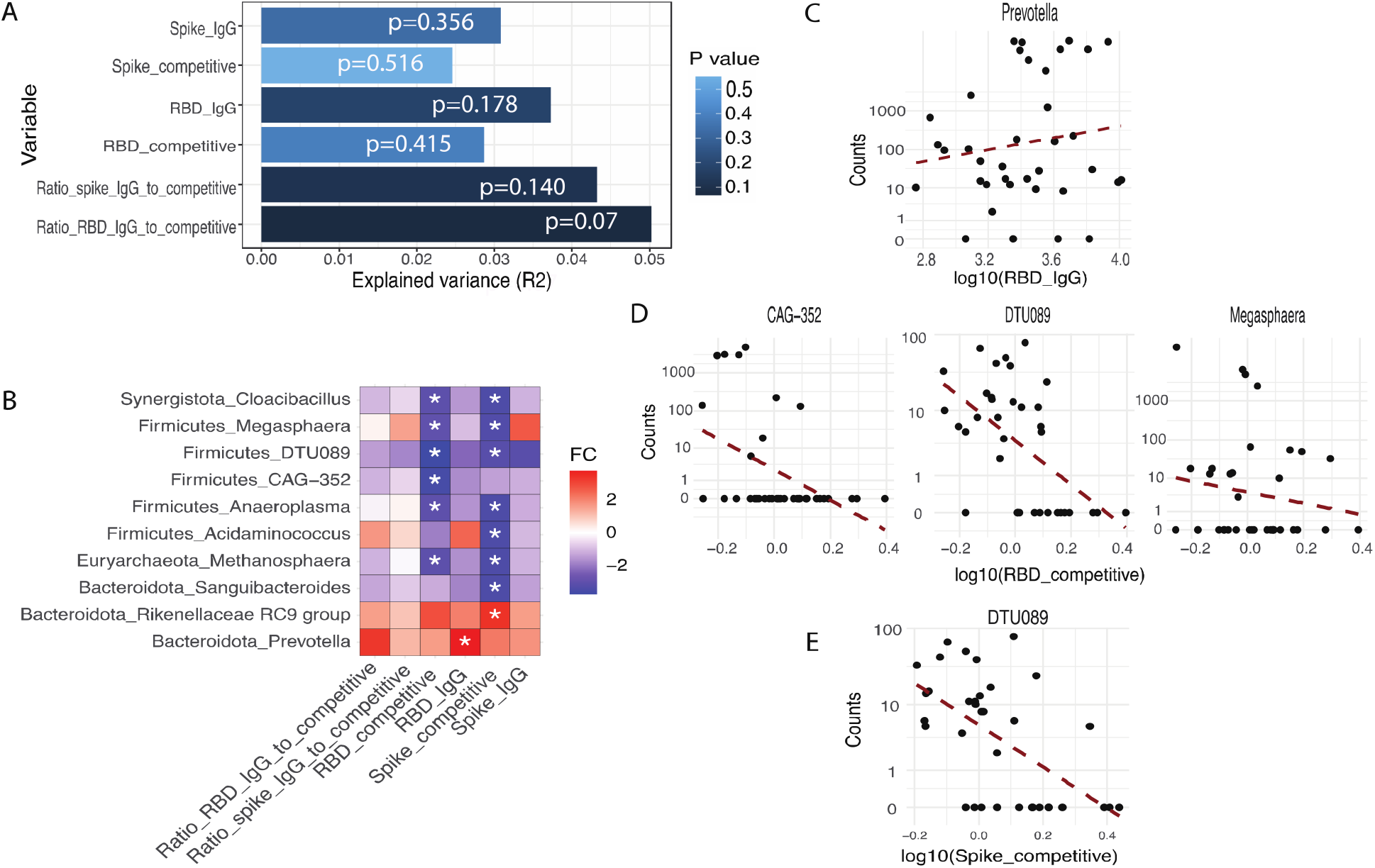
Correlation between baseline gut microbiota taxa and immune parameters twelve-weeks post 1^st^ BNT162b2 vaccine. (A) The variability in RBD and spike IgG, competitive binding antibody and IgG to competitive binding antibody ratio explained by the variability in gut microbiota composition (PERMANOVA). (B) Heat-map of the gut microbiota taxa positively (red) or negatively (blue) correlated with each of the immune parameters (*p<0.05; Wald test). (C) Baseline Prevotella counts were positively associated with RBD IgG levels post 1^st^ BNT162b2 vaccine (p=0.032; Wald test). (D) Baseline *Clostridiales* bacterium DTU089 and CAG-352, and *Megasphaera* counts were negatively associated with RBD competitive binding antibody levels post 1^st^ BNT162b2 vaccine (p=0.038, p=0.038 and p=0.047, respectively; Wald test). (E) Baseline *Clostridiales* bacterium DTU089 counts were negatively associated with spike competitive binding antibody levels post 1^st^ BNT162b2 vaccine (p=0.029; Wald test).

As the functional capacity of the gut microbiome appears to be an important factor in modulating immune responses^19^, we examined whether specific microbial metabolites produced via bacterial fermentation, i.e., SCFAs, were associated with key immune parameters post 1^st^ BTN162b2 vaccine dose. The SCFAs acetate, propionate and butyrate exert anti-inflammatory effects, whereas, relatively little is known about the impact that branched-chain fatty acids (BCFA), such as isobutyric and isovaleric acids, have on immune responses^19^. Baseline BCFA were negatively correlated with several immune parameters (**figure 4A**). Specifically, higher baseline isovaleric acid concentrations (μmol/g stool) were negatively associated with RBD IgG (p=0.0303, r= -0.373; Spearman rank) (**figure 4B**), spike IgG to competitive binding antibody ratio (p=0.033, r= -0.367; Spearman rank) (**figure 4C**) and spike IgG (p=0.0265, r= -0.381; Spearman rank) (**figure 4D**). Baseline isobutyric acid concentrations were also negatively associated with spike IgG to competitive binding antibody ratio (p=0.0179, r= -0.406; Spearman rank) (**figure 4C**).

**Figure 4.**
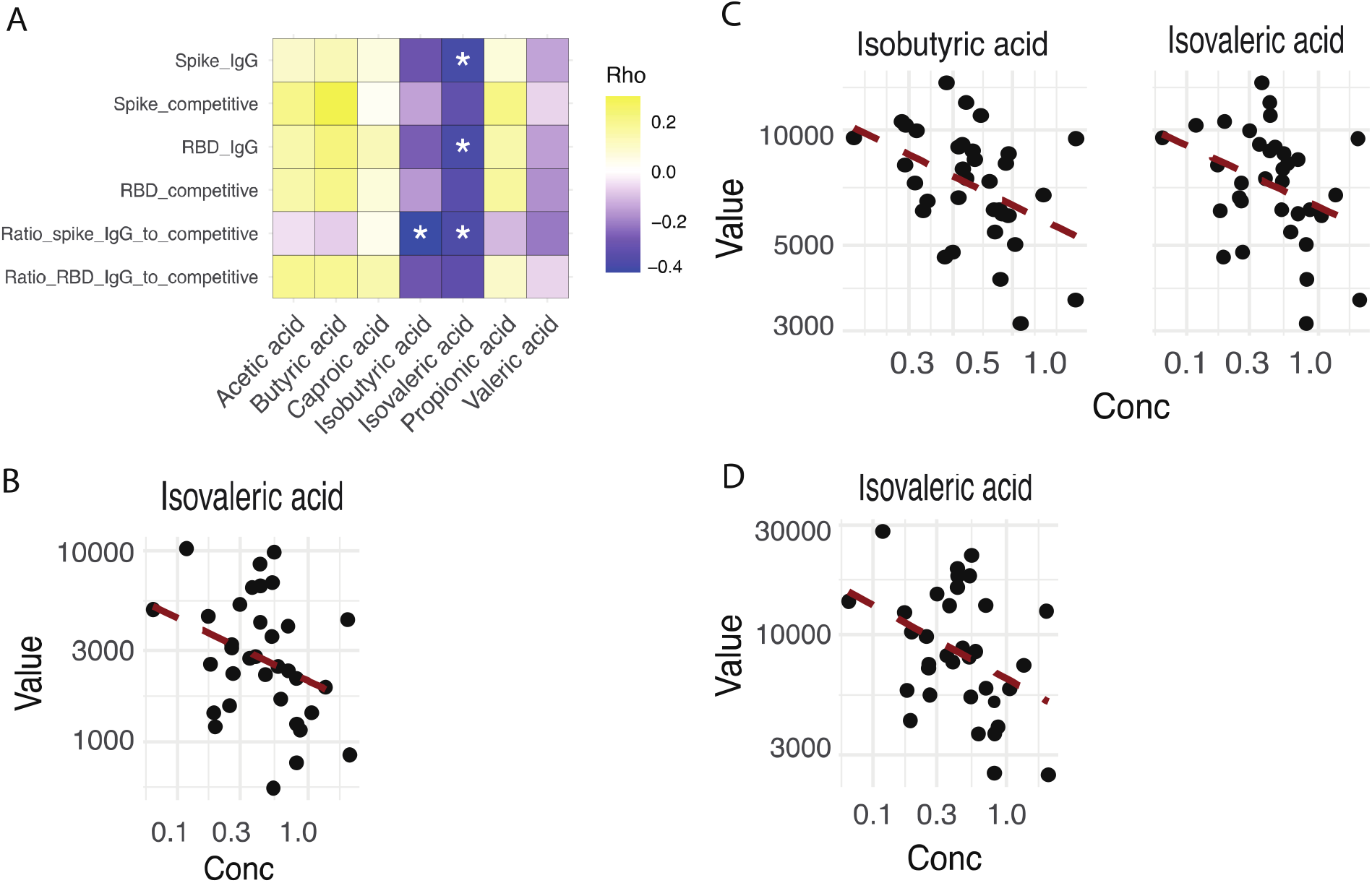
Correlation between baseline SCFA and immune parameters twelve-weeks post 1^st^ BNT162b2 vaccine. (A) Heat-map of the specific SCFAs that were negatively (blue) or positively (yellow) correlated with each of the immune response parameters (*p<0.05; Spearman rank). (B) Baseline isovaleric acid concentrations were negatively associated with RBD IgG levels (p=0.0303, r= -0.373; Spearman rank). (C) Baseline isobutyric and isovaleric acid concentrations were negatively associated with spike IgG to competitive binding antibody ratio levels (p=0.0179, r= -0.406 and p=0.033, r= -0.367, respectively; Spearman rank). (D) Baseline isovaleric acid concentrations were negatively associated with spike IgG levels (p=0.0265, r= -0.381; Spearman rank).

### Baseline gut microbiota and SCFAs associated with responses to BNT162b2 vaccination

As all participants developed a robust immune response to the 1^st^ BNT162b2 vaccine dose, we decided to group participants into quartiles, based on the strength of their response, to determine whether there was a gut microbiota signature that differentiated between low and high vaccine responders.

We observed a trend towards a distinctive gut microbiota signature between low (quartile 1) and high vaccine responders (quartile 4) based on RBD IgG levels (p=0.0554; PERMANOVA) (**figure 5A**). Correspondingly, there were also several baseline bacterial taxa that significantly differed (p<0.05; Wald test) between participants with low versus high BNT162b2 vaccine responses (**figure 5B, figure S4A-D**). The most pronounced bacterial taxa differences were significantly higher baseline counts of *Prevotella* and *Haemophilus* genera in participants with high RBD IgG levels (p=0.00016 and p<0.0001, respectively; Wald test) (**figure 5C, figure S4A**) and RBD IgG to competitive binding antibody ratio (p=0.0003 and p=0.0005, respectively; Wald test) (**figure 5D, figure S4B**). Participants with high RBD competitive binding antibody levels had greater baseline counts of *Haemophilus* and *Veillonella* (p=0.0043 and p<0.0001, respectively; Wald test) (**figure 5E, figure S4C**). Lastly, baseline *Prevotella* counts were also significantly higher in participants with high spike IgG levels (p=0.0054; Wald test) (**figure 5F, figure S4D**). Similar to the study by Ng and colleagues^8^, we also demonstrated that certain baseline gut microbiota are associated with inter-individual responsiveness to the BNT162b2 vaccine. The specific bacterial taxa did, however, differ between our two cohorts with their study identifying that higher abundances of *Eubacterium rectale, Roseburia faces, Bacteroides thetaiotaomicron* and *Bacteroides spp*. OM05-12 were associated with stronger BNT162b2 vaccine responses.

**Figure 5.**
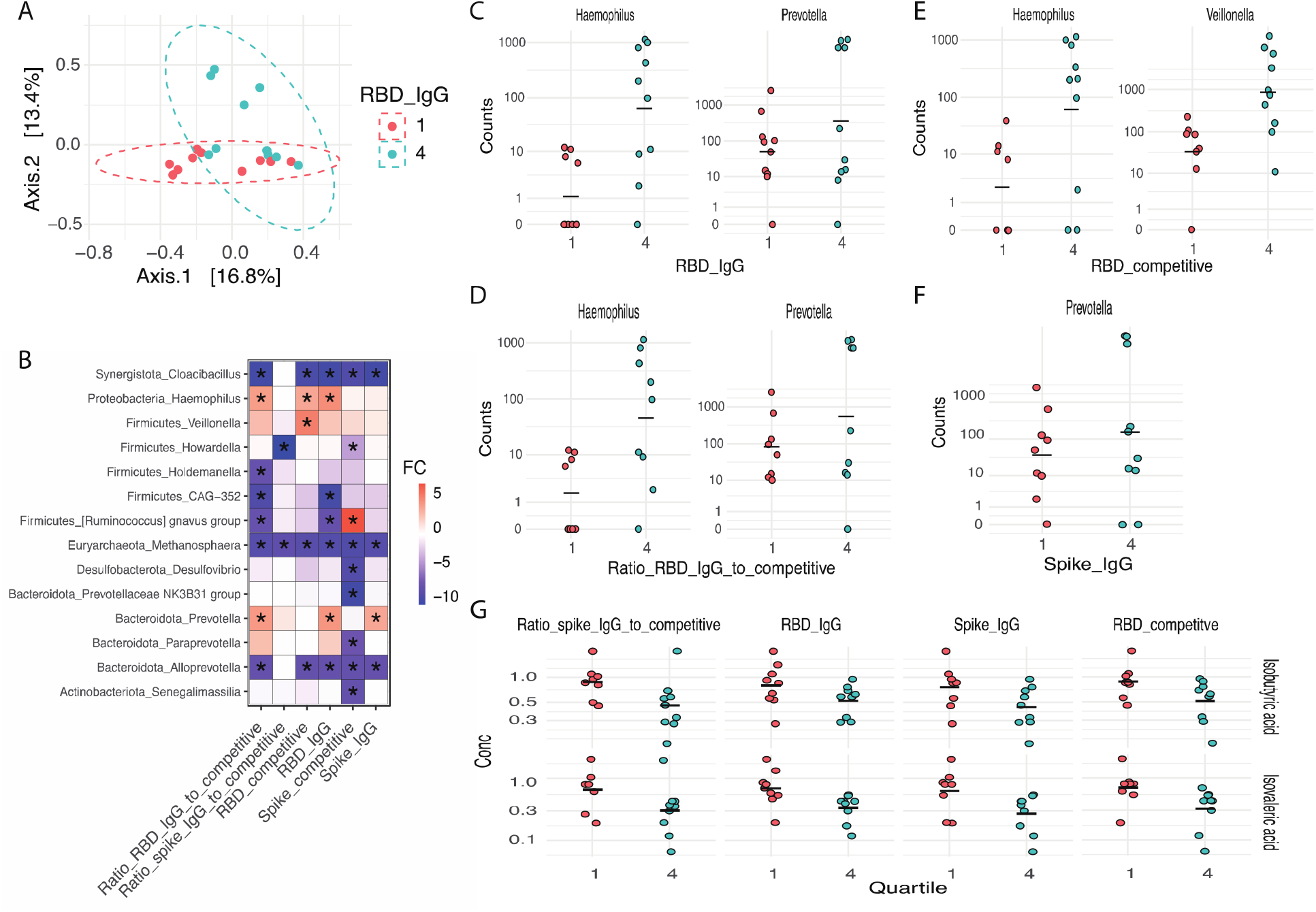
Significant differences in baseline microbial taxa in participants with differential (lowest [quartile 1] versus highest [quartile 4]) immune parameter responses twelve-weeks post 1^st^ BNT162b2 vaccine. (A) A trend towards a significant difference (p=0.0554; PERMANOVA) in the gut microbiota community was observed in participants with the lowest (quartile 1 [pink]) versus the highest (quartile 4 [green]) RBD IgG response twelve-weeks post 1^st^ BNT162b2 vaccine dose. (B) Heat-map depicting several significantly higher (red) or lower (blue) (*p<0.05; Wald test) baseline microbial counts in high (quartile 4) versus low (quartile 1) BTN162b2 vaccine responders across several immune parameters. (C) Baseline Prevotella and Haemophilus counts were significantly (p=0.00016 and p<0.0001, respectively; Wald test) higher in participants with the highest (quartile 4) RBD IgG levels twelve-weeks post 1^st^ BNT162b2 vaccine. (D) Baseline *Prevotella* and *Haemophilus* counts were significantly (p=0.00029 and p=0.00049, respectively; Wald test) higher in participants with the highest (quartile 4) ratio of RBD IgG to competitive binding antibody levels twelve-weeks post 1^st^ BNT162b2 vaccine. (E) Baseline *Haemophilus* and *Veillonella* counts were significantly (p=0.0043 and p<0.0001, respectively; Wald test) higher in participants with the highest (quartile 4) RBD competitive binding antibody levels twelve-weeks post 1^st^ BNT162b2 vaccine. (F) Baseline *Prevotella* counts were significantly (p=0.00544; Wald test) higher in participants with the highest (quartile 4) ratio of RBD IgG to competitive binding antibody twelve-weeks post 1^st^ BNT162b2 vaccine. (G) Individuals with the highest baseline concentrations (μmol/g stool) of isobutyric acid had the lowest spike IgG to competitive binding antibody ratio levels (p=0.045; t-test, Bonferroni corrected) while those with the highest baseline concentrations of isovaleric acid had the lowest RBD competitive binding antibody, RBD IgG and spike IgG levels (p=0.021, p=0.044 and p=0.042, respectively; t-test, Bonferroni corrected).

In addition, we explored whether SCFAs were associated with varying BNT162b2 vaccine responses. In our cohort, we observed significant differences in vaccine-dependent immune responses in participants with distinctive baseline SCFA profiles; specifically, the BCFAs isobutyric and isovaleric acids. Participants with high baseline concentrations (μmol/g stool) of isobutyric acid had the lowest spike IgG to competitive binding antibody ratio (p=0.045; t-test, Bonferroni corrected). Additionally, participants with high baseline concentrations of isovaleric acid had the lowest RBD competitive binding antibody, RBD IgG and spike IgG levels (p=0.021, p=0.044 and p=0.042, respectively; t-test, Bonferroni corrected) (**figure 5G, figure S5**).

### Anti-SARS-CoV-2 spike total relative fractional avidity was significantly associated with several gut microbiota taxa but not SCFA concentrations

IgG avidity maturation has been shown to modulate antibody-mediated functions and vaccine effectiveness in a RTS,S/ AS01E malaria vaccine model and is known to evolve months following SARS-CoV-2 mRNA vaccination^20–22^. To circumvent the rigidity of traditional ELISA-based chaotrope avidity assays we performed a multi-point titration of ammonium thiocyanate to measure the quantity of very low to high avidity serum IgG by increasing concentrations of chaotrope^11,12^. Specifically, we assessed the avidity of BNT162b2 vaccine-elicited IgG to determine whether the strength of antibody binding was associated with a specific gut microbiome signature in the subset of participants in which blood was collected twelve weeks after the 1^st^ dose (timepoint V2) as well as four weeks after the 2^nd^ vaccine dose (timepoint V3; n=15). As expected, we found that the avidity at timepoint V3 was significantly higher than at V2 (data not shown).

SCFA concentrations at baseline or twelve-weeks post 1^st^ BNT162b2 vaccine dose were not significantly associated with total relative fractional avidity (TRFA) (**figure S6**). However, compositional features of the gut microbiota were significantly associated with TRFA after the 1^st^ BNT162b2 vaccine dose (p=0.0438, r=0.094; Spearman rank; V2). Moreover, several bacterial species were significantly correlated with TRFA after the 1^st^ (V2) and 2^nd^ (V3) BNT162b2 vaccine dose. Specifically, higher TRFA after V2 was significantly associated with higher baseline abundance of *Slackia isoflavoniconvertens, Phascolarctobacterium succinatutens, Howardella ureilytica, Bacteroides plebeius, Bacteroides stercoris* and *Alistipes inops* (p<0.01; Wald test, Benjamini-Hochberg adjusted) (**figure 6; table S2**). Additionally, lower baseline abundances of *Sutterella wadsworthensis, Lachnospiraceae UCG-001 bacterium, Bacteroides coprocola* and *Acidaminococcus intestini* (p<0.01; Wald test, Benjamini-Hochberg adjusted) were associated with higher TRFA after V2 (**figure 6, table S2**).

**Figure 6.**
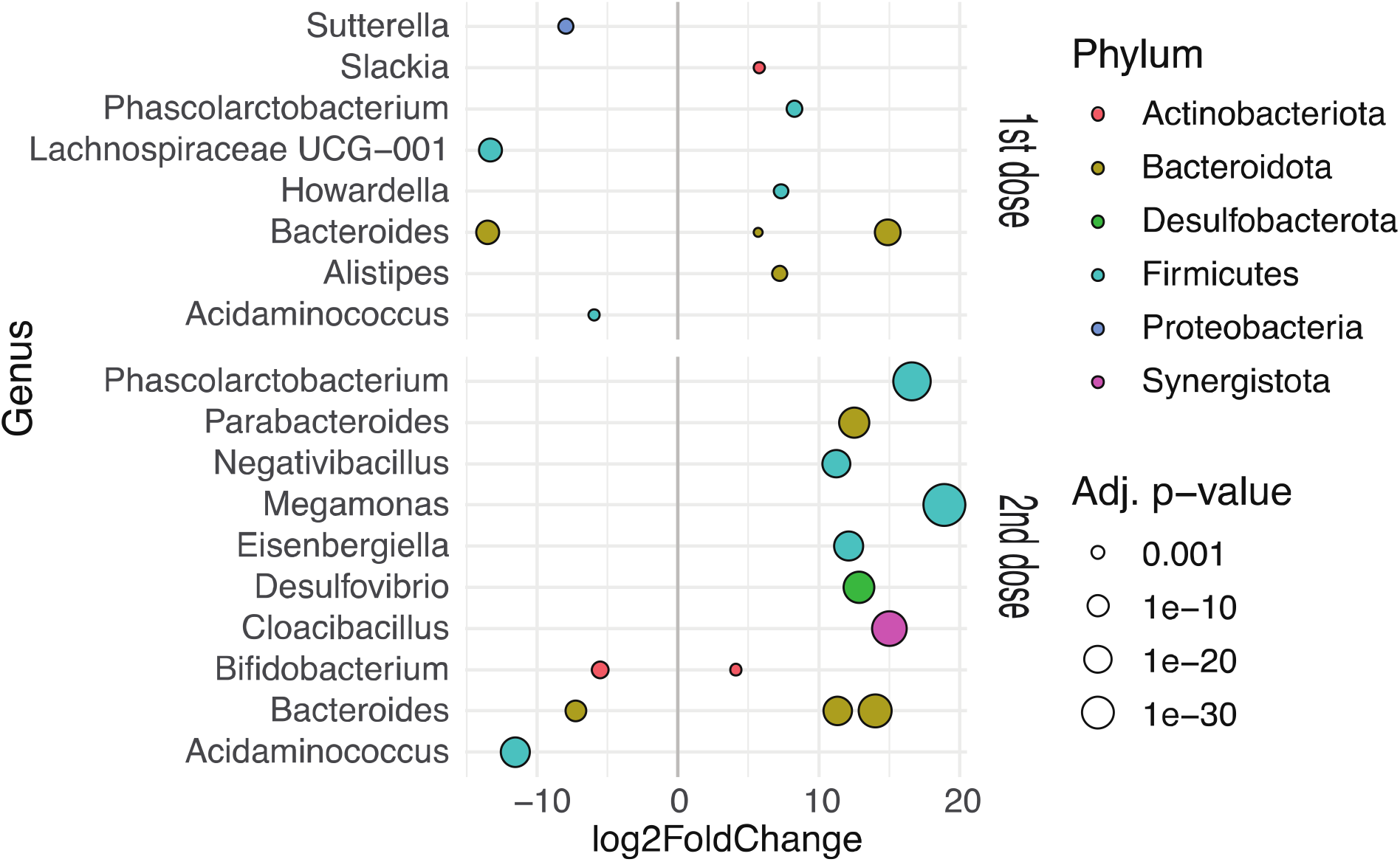
Significant associations between certain microbial species and total relative fractional avidity (TRFA). Several gut microbiota species were significantly positively or negatively associated (p<0.01; Wald test) with higher avidity after the 1^st^ dose of the BNT162b2 vaccine (V2) in a subset of participants (n=15). Several gut microbiota species were also significantly positively or negatively associated (p<0.01; Wald test) with higher avidity after the 2^nd^ dose of the BNT162b2 vaccine (V3) in a subset of participants (n=15). Refer to table 3 and 4 for the specific species names and p values.

After V3 there were several microbial species that were also positively associated with higher TRFA including *Phascolarctobacterium succinatutens, Parabacteroides johnsonii, Negativibacillus massilliensis, Megamonas funiformis, Eisenbergiella tayi, Desulfovibrio fairfieldensis, Cloacibacillus evryensis, Bifidobacterium animalis, Bacteroides plebeius and Bacteroides ovatus* (p<0.01; Wald test, Benjamini-Hochberg adjusted) (**figure 6; table S3**). Lastly, lower baseline abundances of Bifidobacterium bifidum, *Bacteroides* massilliensis and *Acidaminococcus intestini* (p<0.0001; Wald test, Benjamini-Hochberg adjusted) were associated with higher TRFA after V3 (**figure 6; table S3**). These data suggest that a baseline gut microbiota signature could predict the functional binding of BNT162b2 vaccine antibody responses between individuals.

### Habitual dietary fibre intakes lead to differences in gut microbiota and total relative fractional avidity to the BNT162b2 vaccine

Previous studies have suggested that distinct habitual dietary intakes could lead to differing gut microbiota responses to various therapies^23^. No past research has elucidated whether habitual dietary intakes, including fibre, impact SARS-CoV-2 vaccine responses via the gut microbiota. Participants were classified as low (males <22 g/day, females <18 g/day), moderate (males 22 to 29.9 g/day, females 18 to 24.9 g/day), or high (males ≥30 g/day, females ≥25 g/day) dietary fibre consumers using a validated habitual dietary fibre intake food frequency questionnaire^24^. There were trends towards a significant difference in microbial community structure between low, moderate, and high dietary fibre consumers at baseline (**figure 7A**) and twelve-weeks post 1^st^ dose (**figure 7A**) of the BNT162b2 vaccine (p=0.061 and p=0.0558, respectively; PERMANOVA). Additionally, upon separating participants into low, moderate, and high dietary fibre consumer groups we observed that the gut microbiome had shifted significantly by twelve-weeks post 1^st^ BNT162b2 vaccine dose in those individuals that were low dietary fibre consumers (p=0.0332; PERMANOVA) but a similar effect was not seen in moderate or high dietary fibre consumers (p=0.511 and p=0.517, respectively; PERMANOVA) (**figure 7B**). Low dietary fibre consumers may, therefore, be more prone to alterations in their gut microbiota post BNT162b2 vaccination.

**Figure 7.**
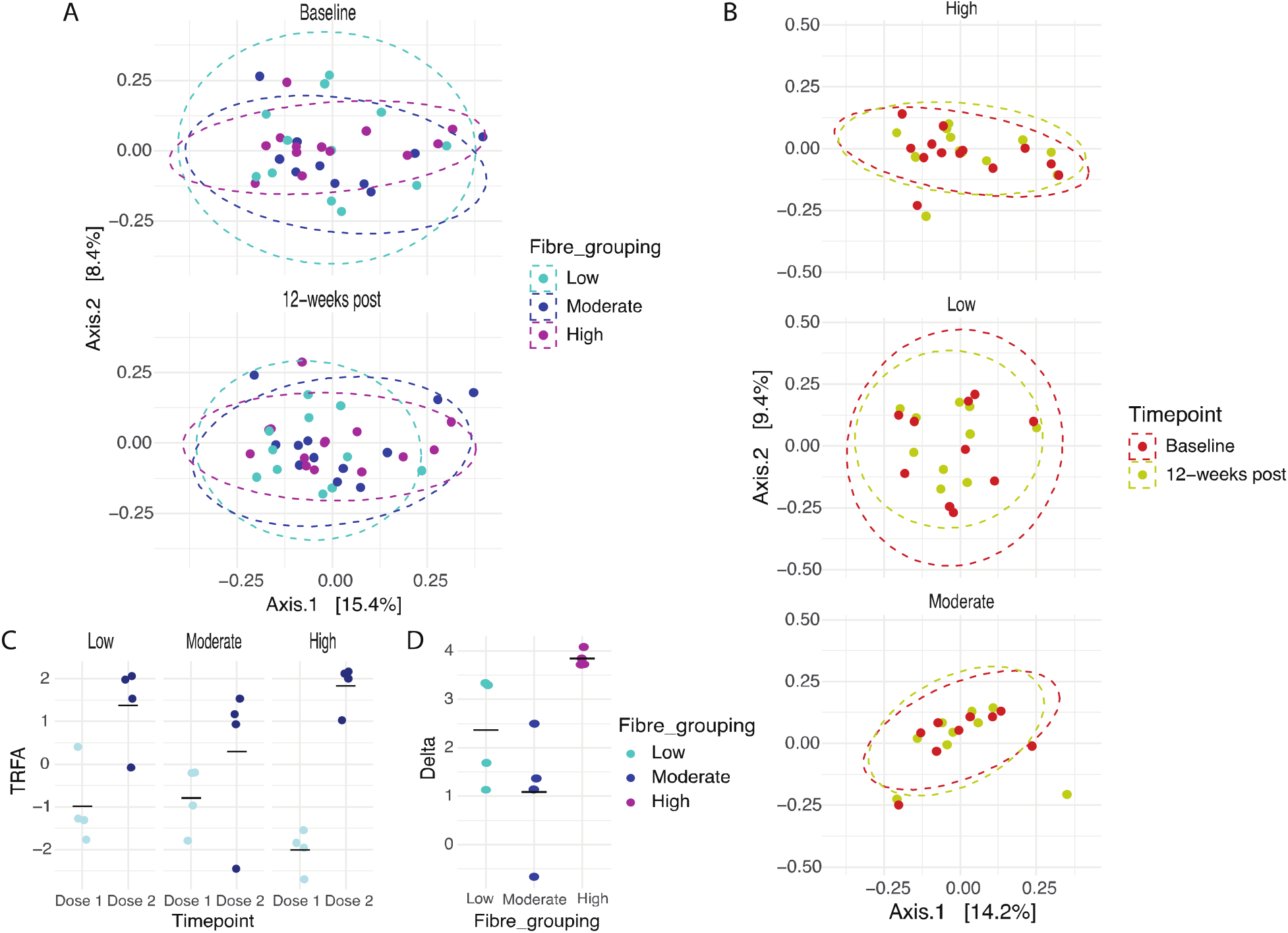
Differences in the gut microbiota community between low, moderate, and high habitual dietary fibre consumers at baseline and twelve-weeks post 1^st^ dose of the BNT162b2 vaccine. (A) At baseline, differing habitual dietary fibre intakes led to a trend towards significant differences (p=0.061; PERMANOVA) in the structure of the gut microbiota community. Twelve-weeks post 1^st^ dose of the BNT162b2 vaccine differing habitual dietary fibre intakes led to a trend towards significant differences (p=0.0558; PERMANOVA) in the structure of the gut microbiota community. (B) A significant shift in the gut microbiota was revealed in low (p=0.0332; PERMANOVA) but not moderate and high dietary fibre consumers (p=0.511 and p=0.517, respectively; PERMANOVA) twelve-weeks after the 1^st^ dose of the BNT162b2 vaccine. (C) After the 1^st^ dose of the BNT162b2 vaccine TRFA was lower in high dietary fibre consumers but not significantly (p=0.058; Kruskal-Wallis). (D) High dietary fibre consumers had a significantly (p=0.018; Kruskal-Wallis) greater change in TRFA from the 1^st^ to 2^nd^ BNT162b2 dose compared to low and moderate dietary fibre consumers.

When comparing the association of differing habitual dietary fibre intakes with immune response twelve weeks after the 1^st^ BTN162b2 vaccine dose, high dietary fibre consumers appeared to have lower TRFA compared to low and moderate dietary fibre consumers, though this did not reach significance (p=0.058; Kruskal-Wallis, **figure 7C**). Interestingly, following the 2^nd^ BTN162b2 dose TRFA values were more similar amongst the groups and less dependent on dietary fibre intake. In addition, the increase in TRFA between doses was more pronounced in high dietary fibre consumers (p=0.018; Kruskal-Wallis, **figure 7D**), suggesting that differing habitual diets may impact the change in IgG binding strength.

## DISCUSSION

Even though SARS-CoV-2 vaccines are highly effective, many vaccinated people still get infected by SARS-Cov-2, with subpopulations developing severe disease^25^. Generating sufficient knowledge of the factors underlying COVID-19 vaccine immunogenicity is therefore important. The multitude of microorganisms that reside within our gut are known to facilitate proper development of the immune system, with an imbalance in gut microbiota, i.e., dysbiosis, implicated in impaired immune responses^26^. Recent studies also highlight the important crosstalk that exists between the gut and lungs (gut-lung axis) in which the gut microbiome plays a role in lung health and disease^27^. Nevertheless, the impact of the microbiota on immunity to SARS-CoV-2 vaccination remains poorly characterized. Here we examined the influence baseline gut microbiota composition and function, as well as habitual dietary fibre intakes, have on BNT162b2 vaccine immunogenicity. Utilizing 16S rRNA sequencing and GC-MS, we profiled the gut microbiome at baseline and twelve weeks following the 1^st^ BTN162b2 dose to establish whether a gut microbiome signature was associated with enhanced vaccine responses.

Participants were grouped as low or high vaccine responders to explore whether features of the gut microbiome were associated with the degree of response. Higher baseline counts of *Prevotella*, and *Haemophilus* were observed in participants with higher RBD IgG levels and RBD IgG to competitive binding antibody ratio. Higher baseline counts of *Prevotella* were also shown to be associated with higher spike IgG levels. Lastly, high counts of *Haemophilus* and *Veillonella* were associated with higher RBD competitive binding antibody levels. Consistent with the results from Ng and colleagues, baseline gut microbiota was also shown to be associated with inter-individual responsiveness to the BNT162b2 vaccine^8^. The specific bacterial taxa that were associated with BNT162b2 vaccine response differed between our two cohorts possibly due to differences in geography (Hong Kong versus Canada), dietary habits and/or microbiome sampling timepoint (1^st^ versus 2^nd^ BNT162b2 vaccine dose). In a study which investigated whether exposure to non-infectious gut microbes shaped pre-existing immunity to SARS-CoV-2 in unexposed individuals, it was discovered that SARS-CoV-2 specific T cells can respond to non-viral microbial peptides, defined cultured commensals (including *Prevotella* spp.) and undefined fecal lysate^28^. As a higher frequency of pre-existing SARS-CoV-2 T cells were associated with milder disease and lower infection rates^29,30^ it is plausible that differences in commensal microbe-based stimulation of SARS-CoV-2 specific immune cells prior to vaccination could impact COVID-19 vaccine responses.

When assessing the antibody binding strength using an avidity assay, we also observed several bacterial taxa that were associated with functional vaccine responses. No studies have previously explored the impact the gut microbiome has on SARS-CoV-2 avidity, however several of the microbes associated with avidity are known to have immunomodulatory properties. Several *Bacteroides* species exhibited positive associations with TRFA in this cohort. Most notably *Bacteroides ovatus* can induce increased production of IgM and IgG antibodies specific to human cancer cells^31^ and can ameliorate lipopolysaccharide-induced inflammation in mice^32^. Another bacterial species with known immune mediating effects, positively associated with TRFA is *Bifidobacterium animalis*. Given as a probiotic *Bifidobacterium animalis* ssp. *lactis* can significantly increase vaccine-specific IgG production after seasonal influenza vaccination^33^. Thus, bacterial species positively associated with TRFA could serve as adjuvants to enhance vaccine immunogenicity to promote longer-term protective effects of the BNT162b2 vaccine. However, this warrants further exploration in a larger cohort.

Another proposed mechanism linking the gut microbiota to vaccine responses is via SCFA dependent B cell production of antibodies^34^ and inhibition of histone deacetylases which regulates the epigenetics of B cells^19^. Altered production of SCFAs, due to dysbiosis, has been shown to disrupt immune responses during COVID-19 infection^35^ suggesting that dysbiosis could also disrupt SARS-CoV-2 vaccine responses. Isovaleric and isobutyric acids are BCFAs which are synthesized exclusively from protein fermentation of branched-chain amino acids^36^. In our cohort, we observed higher isovaleric acid concentrations in participants with low RBD competitive binding antibody, RBD IgG and spike IgG levels. Additionally, higher isobutyric acid concentrations were associated with the lowest spike IgG to competitive binding antibody ratios, suggesting a potentially negative role of BCFAs in vaccine responses. Conversely, other SCFAs such as acetic, propionic, and butyric acids are produced from carbohydrate and protein fermentation^37^. High protein diets lead to higher concentrations of BCFAs, likely due to a shift from carbohydrate to protein utilisation by the gut microbiota^38^. BCFA concentrations have also been shown to be higher in patients with immune-mediated conditions such as inflammatory bowel disease^39^. However, extremely little is known about the mechanisms by which BCFAs modulate inflammatory processes or antibody-mediated vaccine responses. Interestingly, Megasphaera spp., which in our study were also negatively associated with vaccine responses, are prominent isovaleric and isobutyric acid producers^40^. Strong conclusions cannot be drawn from this novel observation, but it highlights the need for future research exploring the impact that BCFAs, and BCFA-producing microbes have on SARS-CoV-2 vaccine responses.

Our study is the first to report an association between habitual dietary intakes and SARS-CoV-2 vaccine immunogenicity. Specifically, low habitual dietary fibre consumers experienced a more pronounced shift in gut microbiota composition after receiving their 1^st^ BNT162b2 vaccine dose. Furthermore, high habitual dietary fibre consumers had lower TRFA after the 1^st^ vaccine dose but a significant increase from the 1^st^ to 2^nd^ dose which was not observed in low or moderate habitual dietary fibre consumers. Unfortunately, stool was not collected after the 2^nd^ vaccine dose, so it is difficult to postulate whether the dramatic increase in TRFA in high habitual dietary fibre consumers was also associated with differential alterations in the gut microbiome as compared to low and moderate consumers. Significant changes in other host responses, i.e., satiety, have been reported in high habitual dietary fibre consumers. Interestingly, these more pronounced host responses were also associated with more significant changes in the gut microbiota after a prebiotic intervention when compared to low habitual dietary fibre consumers^41^. Although researchers have not focused on the effect habitual diet has on vaccine responses, since diet is one of the main modifying factors of the gut microbiota, it is certainly feasible that diet-dependent modulation of the gut microbiota may alter vaccine immunogenicity. Protein deficient diets which lead to microbial dysbiosis can reduce oral attenuated human rotavirus vaccine responses^42^ and dysbiosis in infants with malnutrition has been correlated with lower efficacy of oral polio vaccines^43^. Additionally, specific nutraceutical deficiencies (i.e., vitamin C and D, omega-3s, zinc, selenium, and magnesium) have been proposed to alter the gut microbiota and increase COVID-19 severity^44^. In contrast, the impact dietary fibres, which are fermented by resident microbiota to produce SCFAs, have on COVID-19 severity or vaccine responses is unclear.

In summary, we demonstrate a link between the gut microbiome and individualized immune responses to the BNT162b2 vaccine. In line with the Ng and colleagues’ study, we observed that several bacterial taxa were associated with altered vaccine responses, however, for the first time this study also showed that BCFA levels may negatively impact vaccine responses, while dietary factors, such as fibre intake, may modulate antibody maturation to mRNA vaccination. Currently, there are several studies underway which aim to explore the therapeutic potential of microbiome-targeted interventions in enhancing vaccine immunogenicity^35^. Novel strategies such as these are needed to help enhance and sustain SARS-CoV-2 vaccine responses.

## Data Availability

All data produced in the present study are available upon reasonable request to the authors

## Acknowledgements

We would like to take the opportunity to thank all the study participants for providing stool and blood samples and volunteering their time to our study. We would like to thank Lauren Muttucomoroe, Tisha Montgomery, Maria Laura Munoz and Azita Harriran for helping recruit and collect samples from participants, and the BC Children’s Hospital Research Institute Gut4Health Microbiome Sequencing CORE and the University of British Columbia Sequencing and Bioinformatic Consortium for undertaking 16S rRNA sequencing. We also thank Mr. Roger Dyer for analyzing the stool short-chain fatty acids and the Weston Family Microbiome Initiative for providing the funding to undertake this study.

## Notes

### Competing Interest Statement

The authors have declared no competing interest.

### Funding Statement

This study was funded by Weston Family Microbiome Initiative (Grant ID GR018179)

### Author Declarations

Clinical Research Ethics Board of the University of British Columbia gave ethical approval for this work

## REFERENCES

1. Patel R, Kaki M, Potluri VS, Kahar P, Khanna D. A comprehensive review of SARS-CoV-2 vaccines: Pfizer, Moderna & Johnson & Johnson. Hum Vaccines Immunother. 2022;18(1). doi:10.1080/21645515.2021.2002083

2. Amodio E, Capra G, Casuccio A, et al. Antibodies responses to SARS-CoV-2 in a large cohort of vaccinated subjects and seropositive patients. Vaccines. 2021;9(7). doi:10.3390/vaccines9070714

3. Chivu-Economescu M, Bleotu C, Grancea C, et al. Kinetics and persistence of cellular and humoral immune responses to SARS-CoV-2 vaccine in healthcare workers with or without prior COVID-19. J Cell Mol Med. 2022;26(4):1293-1305. doi:10.1111/jcmm.17186

4. Levine-Tiefenbrun M, Yelin I, Alapi H, et al. Waning of SARS-CoV-2 booster viral-load reduction effectiveness. Nat Commun. 2022;13(1):1–4. doi:10.1038/s41467-022-28936-y

5. Collier DA, Ferreira IATM, Kotagiri P, et al. Age-related immune response heterogeneity to SARS-CoV-2 vaccine BNT162b2. Nature. 2021;596(7872):417–422. doi:10.1038/s41586-021-03739-1

6. Grupper A, Rabinowich L, Schwartz D, et al. Reduced humoral response to mRNA SARS-CoV-2 BNT162b2 vaccine in kidney transplant recipients without prior exposure to the virus. Am J Transplant. 2021;21(8):2719–2726. doi:10.1111/ajt.16615

7. Madison AA, Shrout MR, Renna ME, Kiecolt-Glaser JK. Psychological and behavioral predictors of vaccine efficacy: Considerations for COVID-19. Perspect Psychol Sci. 2021;16(2):191–203. doi:10.1177/1745691621989243

8. Ng SC, Peng Y, Zhang L, et al. Gut microbiota composition is associated with SARS-CoV-2 vaccine immunogenicity and adverse events. Gut. 2022;(January):gutjnl-2021-326563. doi:10.1136/gutjnl-2021-326563

9. Frej-Mądrzak M, Jeziorek M, Sarowska J, Jama-Kmiecik A, Choroszy-Król I. The role of probiotics and prebiotics in the proper functioning of gut microbiota and the treatment of diseases caused by gut microbiota dysbiosis. Nutr Obes Metab Surg. 2020;7(1):9–15. doi:10.5114/noms.2020.94667

10. Johnson M, Wagstaffe HR, Gilmour KC, et al. Evaluation of a novel multiplexed assay for determining IgG levels and functional activity to SARS-CoV-2. J Clin Virol. 2020;130:104572. doi:10.1016/j.jcv.2020.104572

11. Abu Raya B, Bamberger E, Almog M, Peri R, Srugo I, Kessel A. Immunization of pregnant women against pertussis: The effect of timing on antibody avidity. Vaccine. 2015;33(16):1948–1952. doi:10.1016/J.VACCINE.2015.02.059

12. Abu-Raya B, Giles ML, Kollmann TR, Sadarangani M. Profiling avidity of antibodies elicited by vaccination using enzyme-linked immunosorbent assay-based elution – Insights into a novel experimental and analytical approach. Vaccine. 2020;38(34):5389–5392. doi:10.1016/j.vaccine.2020.06.060

13. Kozich JJ, Westcott SL, Baxter NT, Highlander SK, Schloss PD. Development of a dual-index sequencing strategy and curation pipeline for analyzing amplicon sequence data on the miseq illumina sequencing platform. Appl Environ Microbiol. 2013;79(17):5112–5120. doi:10.1128/AEM.01043-13

14. Laidlaw BJ, Ellebedy AH. The germinal centre B cell response to SARS-CoV-2. Nat Rev Immunol. 2022;22(1):7–18. doi:10.1038/s41577-021-00657-1

15. Bartsch YC, Fischinger S, Siddiqui SM, et al. Discrete SARS-CoV-2 antibody titers track with functional humoral stability. Nat Commun 2021 121. 2021;12(1):1–8. doi:10.1038/s41467-021-21336-8

16. Thomas AS, Coote C, Moreau Y, et al. Antibody-dependent cellular cytotoxicity responses and susceptibility influence HIV-1 mother-to-child transmission. JCI Insight. 2022;7(9). doi:10.1172/JCI.INSIGHT.159435

17. Jarlhelt I, Nielsen SK, Jahn CXH, et al. SARS-CoV-2 Antibodies Mediate Complement and Cellular Driven Inflammation. Front Immunol. 2021;12:4612. doi:10.3389/FIMMU.2021.767981/BIBTEX

18. Sancilio A, D’aquila R, Mcnally EM, et al. A surrogate virus neutralization test to quantify antibody-mediated inhibition of SARS-CoV-2 in finger stick dried blood spot samples. medRxiv. February 2021:2021.02.14.21251709. doi:10.1101/2021.02.14.21251709

19. Yao Y, Cai X, Fei W, Ye Y, Zhao M, Zheng C. The role of short-chain fatty acids in immunity, inflammation and metabolism. Crit Rev Food Sci Nutr. 2022;62(1):1–12. doi:10.1080/10408398.2020.1854675

20. Tauzin A, Gong SY, Beaudoin-Bussières G, et al. Strong humoral immune responses against SARS-CoV-2 Spike after BNT162b2 mRNA vaccination with a 16-week interval between doses. Cell Host Microbe. December 2021. doi:10.1016/J.CHOM.2021.12.004

21. Dobaño C, Sanz H, Sorgho H, et al. Concentration and avidity of antibodies to different circumsporozoite epitopes correlate with RTS,S/AS01E malaria vaccine efficacy. Nat Commun 2019 101. 2019;10(1):1–13. doi:10.1038/s41467-019-10195-z

22. Thompson HA, Hogan AB, Walker PGT, et al. Modelling the roles of antibody titre and avidity in protection from Plasmodium falciparum malaria infection following RTS,S/AS01 vaccination. Vaccine. 2020;38(47):7498–7507. doi:10.1016/J.VACCINE.2020.09.069

23. Healey GR, Murphy R, Brough L, Butts CA, Coad J. Interindividual variability in gut microbiota and host response to dietary interventions. Nutr Rev. 2017;75(12):1059–1080. doi:10.1093/nutrit/nux062

24. Healey G, Brough L, Murphy R, Hedderley D, Butts C, Coad J. Validity and reproducibility of a habitual dietary fibre intake short food frequency questionnaire. Nutrients. 2016;8(9):3–9. doi:10.3390/nu8090558

25. Tenforde MW, Self WH, Adams K, et al. Association between mRNA vaccination and COVID-19 hospitalization and disease severity. J Am Med Assoc. 2021;326(20):2043–2054. doi:10.1001/jama.2021.19499

26. Zheng D, Liwinski T, Elinav E. Interaction between microbiota and immunity in health and disease. Cell Res. 2020;30(6):492–506. doi:10.1038/s41422-020-0332-7

27. Dang AT, Marsland BJ. Microbes, metabolites, and the gut–lung axis. Mucosal Immunol. 2019;12(4):843–850. doi:10.1038/s41385-019-0160-6

28. Bartolo L, Afroz S, Pan Y-G, et al. SARS-CoV-2-specific T cells in unexposed adults display broad trafficking potential and cross-react with commensal antigens. Sci Immunol. 2022;3127. https://www.biorxiv.org/content/10.1101/2021.11.29.470421v1 https://www.biorxiv.org/content/10.1101/2021.11.29.470421v1.abstract.

29. Mallajosyula V, Ganjavi C, Chakraborty S, et al. CD8+T cells specific for conserved coronavirus epitopes correlate with milder disease in COVID-19 patients. Sci Immunol. 2021;6(61). doi:10.1126/sciimmunol.abg5669

30. Swadling L, Diniz MO, Schmidt NM, et al. Pre-Existing Polymerase-Specific T Cells Expand in Abortive Seronegative SARS-CoV-2. Vol 601.; 2021. doi:10.1038/s41586-021-04186-8

31. Ulsemer P, Henderson G, Toutounian K, et al. Specific humoral immune response to the Thomsen-Friedenreich tumor antigen (CD176) in mice after vaccination with the commensal bacterium Bacteroides ovatus D-6. Cancer Immunol Immunother. 2013;62(5):875–887. doi:10.1007/s00262-013-1394-x

32. Tan H, Zhao J, Zhang H, Zhai Q, Chen W. Novel strains of Bacteroides fragilis and Bacteroides ovatus alleviate the LPS-induced inflammation in mice. Appl Microbiol Biotechnol. 2019;103(5):2353–2365. doi:10.1007/s00253-019-09617-1

33. Rizzardini G, Eskesen D, Calder PC, Capetti A, Jespersen L, Clerici M. Evaluation of the immune benefits of two probiotic strains Bifidobacterium animalis ssp. lactis, BB-12 ® and Lactobacillus paracasei ssp. paracasei, L. casei 431 ® in an influenza vaccination model: A randomised, double-blind, placebo-controlled study. Br J Nutr. 2012;107(6):876–884. doi:10.1017/S000711451100420X

34. Yamamoto I, Adachi T, Kishiro Y, Fujiwara M, Gohda E. Interleukin-2 - dependent augmentation of the anti-tnp antibody production by sodium butyrate in cultured murine splenic B cells. Int J Immunopharmacol. 1997;19(6):347–354.

35. Chen J, Vitetta L, Henson JD, Hall S. The intestinal microbiota and improving the efficacy of COVID-19 vaccinations. J Funct Foods. 2021;87:104850. doi:10.1016/j.jff.2021.104850

36. Smith EA, Macfarlane GT. Dissimilatory amino acid metabolism in human colonic bacteria. Anaerobe. 1997;3(5):327–337. doi:10.1006/anae.1997.0121

37. Gilbert MS, Ijssennagger N, Kies AK, van Mil SWC. Protein fermentation in the gut; implications for intestinal dysfunction in humans, pigs, and poultry. Am J Physiol - Gastrointest Liver Physiol. 2018;315(2):G159–G170. doi:10.1152/ajpgi.00319.2017

38. Russell WR, Gratz SW, Duncan SH, et al. High-protein, reduced-carbohydrate weight-loss diets promote metabolite profiles likely to be detrimental to colonic health. Am J Clin Nutr. 2011;93(5):1062–1072. doi:10.3945/ajcn.110.002188

39. Van Nuenen MHMC, Venema K, Van Der Woude JCJ, Kuipers EJ. The metabolic activity of fecal microbiota from healthy individuals and patients with inflammatory bowel disease. Dig Dis Sci. 2004;49(3):485–491. doi:10.1023/B:DDAS.0000020508.64440.73

40. Dai Z, Wu G, Zhu W. Amino acid metabolism in intestinal bacteria: links between gut ecology and host health. Front Biosci. 2011;16:1768–1786. doi:10.1109/leoswt.2008.4444364

41. Healey G, Murphy R, Butts C, Brough L, Whelan K, Coad J. Habitual dietary fibre intake influences gut microbiota response to an inulin-type fructan prebiotic: a randomised, double-blind, placebo-controlled, cross-over, human intervention study. Br J Nutr. 2018. doi:10.1017/S0007114517003440

42. Kumar A, Vlasova AN, Deblais L, et al. Impact of nutrition and rotavirus infection on the infant gut microbiota in a humanized pig model. BMC Gastroenterol. 2018;18(1):1–17. doi:10.1186/s12876-018-0810-2

43. Haque R, Snider C, Liu Y, et al. Oral polio vaccine response in breast fed infants with malnutrition and diarrhea. Vaccine. 2014;32(4):478–482. doi:10.1016/j.vaccine.2013.11.056

44. Rishi P, Thakur K, Vij S, et al. Diet, gut microbiota and COVID-19. Indian J Microbiol. 2020;60(4):420–429. doi:10.1007/s12088-020-00908-0

