## Supplementary information for "Variability in mRNA SARS-CoV-2 BNT162b2 vaccine immunogenicity is associated with differences in the gut microbiome and habitual dietary fibre intake"

**Supplementary methods**

**Study cohort**

Due to the rapid initiation of SARS-CoV-2 vaccine programs for healthcare workers in British Columbia this research was particularly time sensitive as samples needed to be collected promptly prior to participants receiving their 1^st^ dose of the BNT612b2 vaccine. Therefore, we did not have an opportunity to involve participants in the design or implementation of this study.

**Participant characteristic data collection**

Demographic data (e.g., age, sex) was collected from participants via a secure web-based application (REDCap). A standardized form, which included a validated habitual dietary fibre intake food frequency questionnaire^1^, was used to collect information from participants regarding antibiotic use prior to stool collection, significant changes in body weight or food intake over the previous year and whether participants were vegan or vegetarian. These participant characteristics were captured as they are known to modulate the gut microbiota and could potentially confound study results^2^.

**SARS-CoV-2 spike and RBD IgG multiplex assay**

Briefly, plates were blocked using 5% bovine serum albumin for 30 minutes and then washed. To the plate 50 μl of diluted serum, standards, and controls were added and incubated for 2 hours with shaking (700 rpm) at room temperature. After washing, 50 μl of SULFO-TAG™ anti-human IgG were added and incubated for 1 hour with shaking at room temperature. Plates were washed and, immediately after adding 150 μl of MSD GOLD read buffer, were read using MSD QuickPlex SQ120 plate reader and Methodical Mind software. Concentrations of anti-spike and anti-RBD antibodies (AU/ml) were extrapolated to a standard curve using a 4-parameter logistic regression model. Samples above the limit of detection were re-diluted at a higher dilution and retested until within the detectable range.

**SARS-CoV-2 spike and RBD ACE-2 competition assay**

Plates were blocked, washed, and 25 μl of diluted sample, standard, and controls were added and incubated for 1 hour with shaking (700 rpm) at room temperature. Next, without washing, 25 μl of SULFO-TAG™ ACE-2 calibrator was added and incubated for another 1 hour at room temperature with shaking. After washing, plates were read using MSD GOLD read buffer on MSD QuickPlex SQ120 plate reader and software. Concentrations of competitive binding antibodies (U/mL) were extrapolated to a standard curve using a 4-parameter logistic regression model. Samples above the limit of detection were re-diluted at a higher dilution and retested until within the detectable range. To calculate the ratio of competitive binding antibodies to the total IgG, the concentration of ACE-2 competing antibodies (U/ml) were divided by total IgG for spike and RBD, respectively.

**Anti-SARS-CoV-2 spike absolute and relative fractional avidity assay**

Briefly, a stock concentration of ammonium thiocyanate (98.9% NH_4_SCN, chaotrope; Milipore Sigma, cat # A1479; 4.0 mol/L) was prepared in 1X PBS and serially diluted to working solutions of 2.0, 1.0, 0.75, and 0.50 mol/L. Anti-SARS-CoV-2 spike ELISA plates (Invitrogen, cat # BMS2325) were washed and 6-fold diluted serum, standards, and controls were added with the final dilution in-plate. Plates were incubated for 30 minutes at 37^o^C without shaking then washed before adding chaotrope. Each sample was tested at 0, 0.50, 0.75, 1.0 and 2.0 M in the same plate by adding 100 μl of titrate or 1X PBS. To empty wells 100 μl of 1X PBS was added to prevent drying. Following a 30-minute incubation at 37^o^C, plates were washed into a waste bucket, and 100 μl of anti-human HRP-conjugated IgG detection antibody were added. Plates were incubated for another 30 minutes at 37^o^C, washed, and incubated with 100 μl of 3,3’,5,5’-tetramethylbenzidine (TMB) for 15 minutes in the dark at room temperature. To each well 100 μl of stop solution was added and plates were immediately read at 450nm using Bio-Rad iMark plate reader and MMP version 6.3 software. Concentration extrapolation (U/ml) was performed using the plate reader software and a 4-parameter logistic regression model. Untreated sample wells were used as the concentration of total IgG (U/ml). Values below detection limit were assigned ½ of the lowest back-calculated standard concentration. The limit of detection was determined as 2.5 standard deviations above the mean of lowest back-calculated standard concentration (182 U/ml).

**Calculation of absolute and relative fractional avidity**

Chaotrope concentrations were determined *a priori* during optimization for Pfizer BNT162b2 vaccinated serum. To calculate the absolute fractional avidity (AFA; U/ml), the difference in consecutive chaotrope conditions were obtained by the formula:

$${AFA}_{n}=Y_{n}-Y_{n+1}$$

where *AFA_n_* is the absolute fractional avidity of a given range of chaotrope, and *Y_n_* and *Y_n+1_* are concentrations of antibodies (U/ml) at consecutive chaotrope concentrations. Therefore, *AFA_n_* quantifies the concentration of IgG released between chaotrope concentrations *n* and *n+*1. Where *Y_n_* is the upper limit of chaotrope, the formula takes the form:

$${AFA}_{upper}=Y_{upper}$$

The upper range of chaotrope was determined to be 2.0 M being the concentration eliminating the majority of detectable signal with good separation between vaccination statuses. The chaotrope ranges were described as very low (0 – 0.50M), low (0.5 – 0.75M), medium (0.75 – 1.0M), high (1.0 – 2.0M), and very high (2.0M).

To calculate relative fractional avidity (RFA; %), each AFA was divided by the total IgG levels in the untreated condition (0 mol/L NH_4_SCN) and multiplied by 100%. Samples were retested at adjusted dilution factors if the sum of relative avidity profiles did not equal 100% of total IgG (**table S4**).

To quantify avidity as a single variable, we combined the RFA values taken at each chaotrope concentration using a principal component analysis (PCA), defining total relative fractional avidity (TRFA) using the loadings of PC1 as follows:

TRFA = -0.533(RFA_vlo_) + 0.0269(RFA_lo_) + 0.477(RFA_med_) + 0.492(RFA_hi_) + 0.495(RFA_vhi_)

**Stool collection**

Participants were asked to collect their stool samples within 24 hours of receiving the collection kits. The stool sample collection kits contained two DNA Genotek collection tubes (OM-200 for metagenomics and ME-200 for metabolomics), gloves, toilet liner, alcohol wipes, absorbent paper, and a pre-paid envelope. Stool collected in the OM-200 and ME-200 tubes can be stored at room temperature for 30 and 7 days, respectively. Participants were asked to mail their samples to the lab within 24 hours of stool collection. On arrival, samples were immediately aliquoted and stored at -80^o^C.

**Short-chain fatty acid analysis**

The samples were placed into 20 mL headspace autosample vials and acidified water, and deuterium labeled internal standards were added.  The samples were then heated to 95^o^C while being mixed for 40 minutes. Once an equilibrium was reached, where the SCFA in the gas phase was directly proportional to the SCFA in the liquid mixture, the autosampler needle drew 0.5 mL of the gas phase from the vial and injected the gas into an Agilent 8890 gas chromatograph coupled with an Agilent 7010B mass spectrometer (GC-MS) system. The GC-MS was equipped with a CTC-PAL headspace system and Agilent FATWAX column (0.25mm ID X 0.25um phase thickness and 30m length). The carrier gas used was Helium at a flow rate of 1.25 mL/min and samples were injected with a 10:1 split ratio. The column oven was temperature programmed from 90^o^C to 230^o^C over 18 minutes with baseline separation of all SCFA. Calibration curves utilized authentic standards and deuterium labeled internal standards of acetic, propionic, butyric and caproic acids (Millipore Sigma or CDN isotopes). The GC-MS was operated in SRM mode and regression lines were calculated using quadratic fits with correlation coefficients of 0.995 to 0.9995.

**Microbial gDNA extraction and quantification**

In brief, stool samples, collected in DNA Genotek OM-200 tubes, were thawed on ice, vortexed, and a 250 μl aliquot of sample was transferred to the supplied bead beating tube. The samples were homogenized using a benchtop vortex with bead beating tube adaptor at 2500rpm for 10 minutes. The DNA was eluted in 100 μl of elution buffer. DNA concentrations were quantified using the Quant-iT Picogreen dsDNA kit following manufacturer’s instructions.

**16s rRNA sequencing**

Briefly, the V4 region of the 16S rRNA gene was amplified with barcode primers containing the index sequences using a KAPA HiFi HotStart Real-time PCR Master Mix (Roche). PCR product amplification and concentration was monitored on a Bio-Rad CFT Connect Real-Time PCR system. Amplicon libraries were then purified, normalized, and pooled using the SequalPrep^TM^ normalization plate (Applied Biosystems). The pooled library was further purified with Agencourt AMPure XP system (Beckman Coulter) following the manufacturer’s protocol. Library concentrations were verified using a Qubit^TM^ dsDNA high sensitivity assay kit (Invitrogen). The purified pooled libraries were submitted to the Sequencing and Bioinformatics Consortium at the University of British Columbia (Vancouver, Canada) for QC and sequencing on a single MiSeq v2 flow cell, to generate paired-end 250 bp reads. Raw base call data (bcl) were converted into FastQ format using the bcl2fastq conversion software from Illumina.

The raw 16S rRNA sequences have been deposited in the NCBI Short Read Archive (SRA), accession number is pending.

**Bioinformatics**

Raw .fastq files were processed using a custom script based on the R package DADA2 (version 1.20.0). Quality filtering was performed using the filterAndTrim function under the following criteria: 1) First 10 bp of each read removed; 2) Reads with expected error (EE = sum(10^(-Q/10)) greater than 2 were discarded. Following merging forward and reverse reads and removing chimeric sequences, a table of amplicon sequence variants (ASVs) was generated, with taxonomic assignment using the Silva database (version 138). For all downstream analyses except calculations of alpha diversity, amplicon sequence variants (ASVs) present in less than 5% of the samples were removed.

**Data analyses**

Alpha diversity of the samples was calculated with species richness and the Shannon index of diversity on raw counts of ASVs. To assess changes in alpha diversity between baseline and 12-weeks post vaccine, only participants with a sample from both time points were considered, and a Wilcoxon rank test was used to determine if the difference within participants was significantly less than zero. Overall composition of samples (beta diversity) was calculated by PCoA on a Bray-Curtis distance matrix. DESeq2 was used to identify differentially abundant taxonomic groups, using variance-stabilizing transformed counts, with significance determined by a Wald test, adjusted for multiple-inference using the Benjamini-Hochberg method. Associations between immune response variables and overall composition of the gut microbiota were assessed by PERMANOVA using the R package ADONIS. Associations between short chain fatty acids (SCFAs) and immune response variables was determined using spearman rank correlation analysis, Bonferroni corrected for multiple comparisons. To define participants as responders and non-responders, we binned the immune response variables into quartiles to group participants as the lowest response (quartile 1) or the highest response (quartile 4). Using these quartiles, differentially abundant taxa associated with immune response was assessed in the same method as above.

**References**

1.	Healey G, Brough L, Murphy R, Hedderley D, Butts C, Coad J. Validity and reproducibility of a habitual dietary fibre intake short food frequency questionnaire. *Nutrients*. 2016;8(9). doi:10.3390/nu8090558

2.	Mirzayi C, Renson A, Furlanello C, et al. Reporting guidelines for human microbiome research: the STORMS checklist. *Nat Med*. 2021;27(11):1885-1892. doi:10.1038/s41591-021-01552-x

**Supplementary figures**

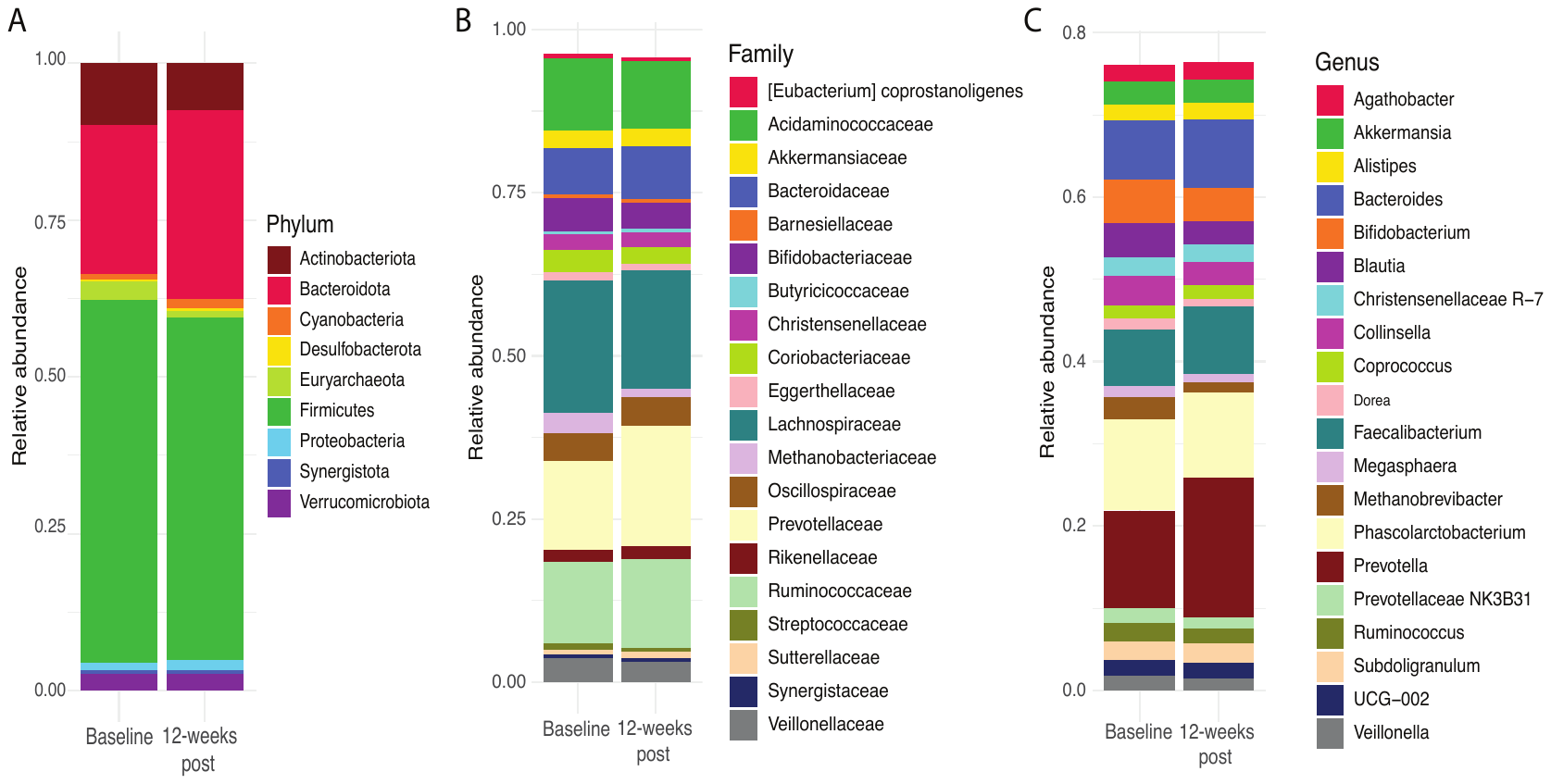

**Supplementary Figure 1** Stacked bar graphs depicting changes in gut microbiota taxonomy from baseline to twelve weeks post 1^st^ BNT162b2 vaccine. (A) Phylum level differences from baseline to twelve weeks post 1^st^ BNT162b2 vaccine. (B) Family level differences from baseline to twelve weeks post 1^st^ BNT162b2 vaccine. (C) Genus level differences from baseline to twelve weeks post 1^st^ BNT162b2 vaccine.

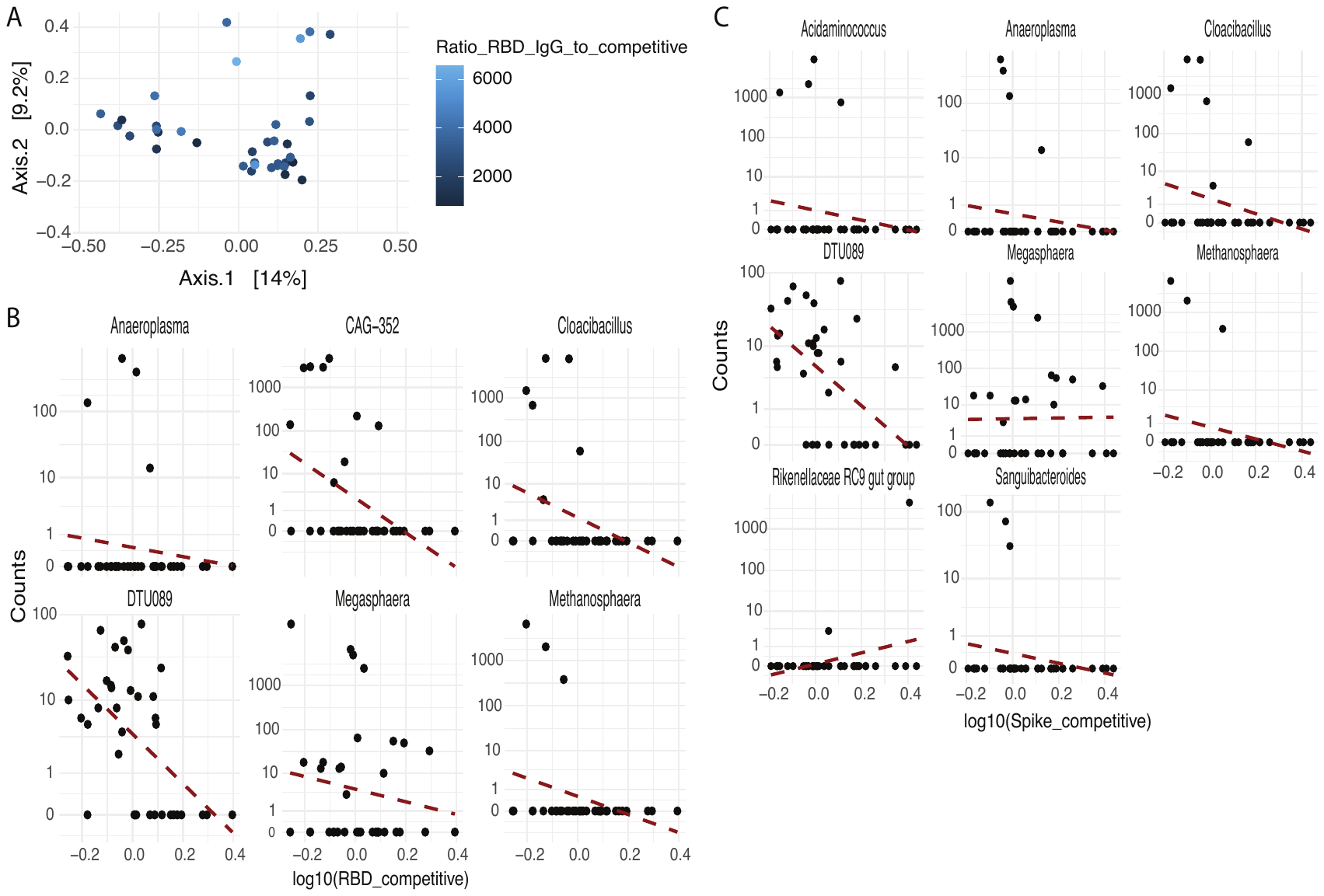

**Supplementary Figure 2** Variability in immune parameters post 1^st^ BNT162b2 vaccine and association with the gut microbiota. (A) Principal co-ordinate analysis plot depicting the individual levels of RBD IgG to competitive binding antibody (circles with different shades of blue) and their relationship to gut microbiota composition – circles which are closer together are more similar in gut microbiota composition then circles further apart. (B) The significant (p<0.05; Wald test) correlations between RBD competitive binding antibody levels and specific gut microbiota taxa. (C) The significant (p<0.05; Wald test) correlations between spike competitive binding antibody levels and specific gut microbiota taxa.

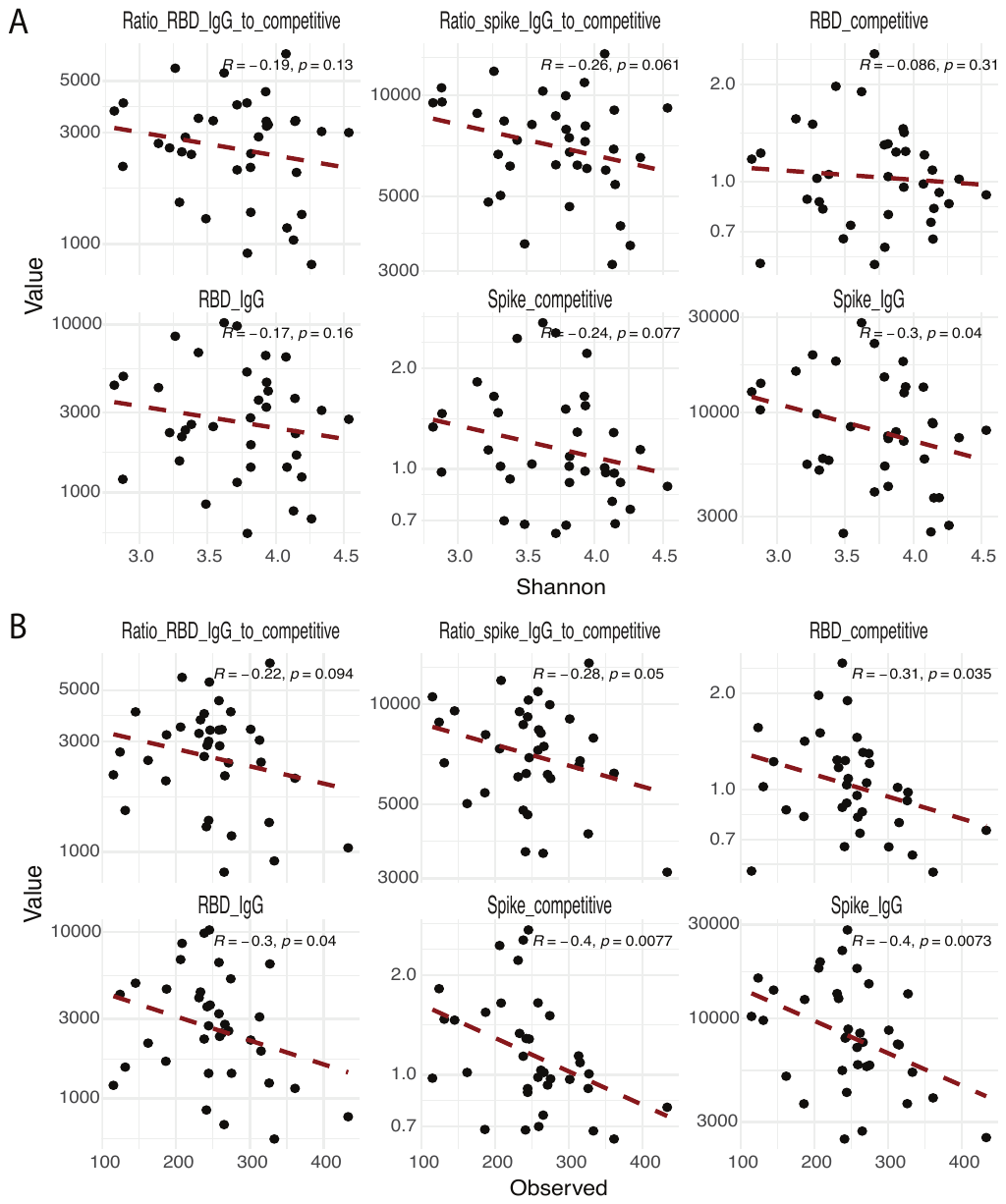

**Supplementary Figure 3** Correlation between baseline alpha diversity and immune parameters twelve-weeks post 1^st^ BNT162b2 vaccine. (A) Shannon alpha diversity was negatively correlated with spike IgG (p=0.04, r= -0.3; Spearman rank) (B) Observed alpha diversity was negatively correlated with RBD competitive binding antibody (p=0.035, r= -0.31; Spearman rank), RBD IgG (p=0.04, r= -0.3; Spearman rank), spike competitive binding antibody (p=0.0077, r= -0.4; Spearman rank) and spike IgG (p=0.0073, r= -0.4; Spearman rank).

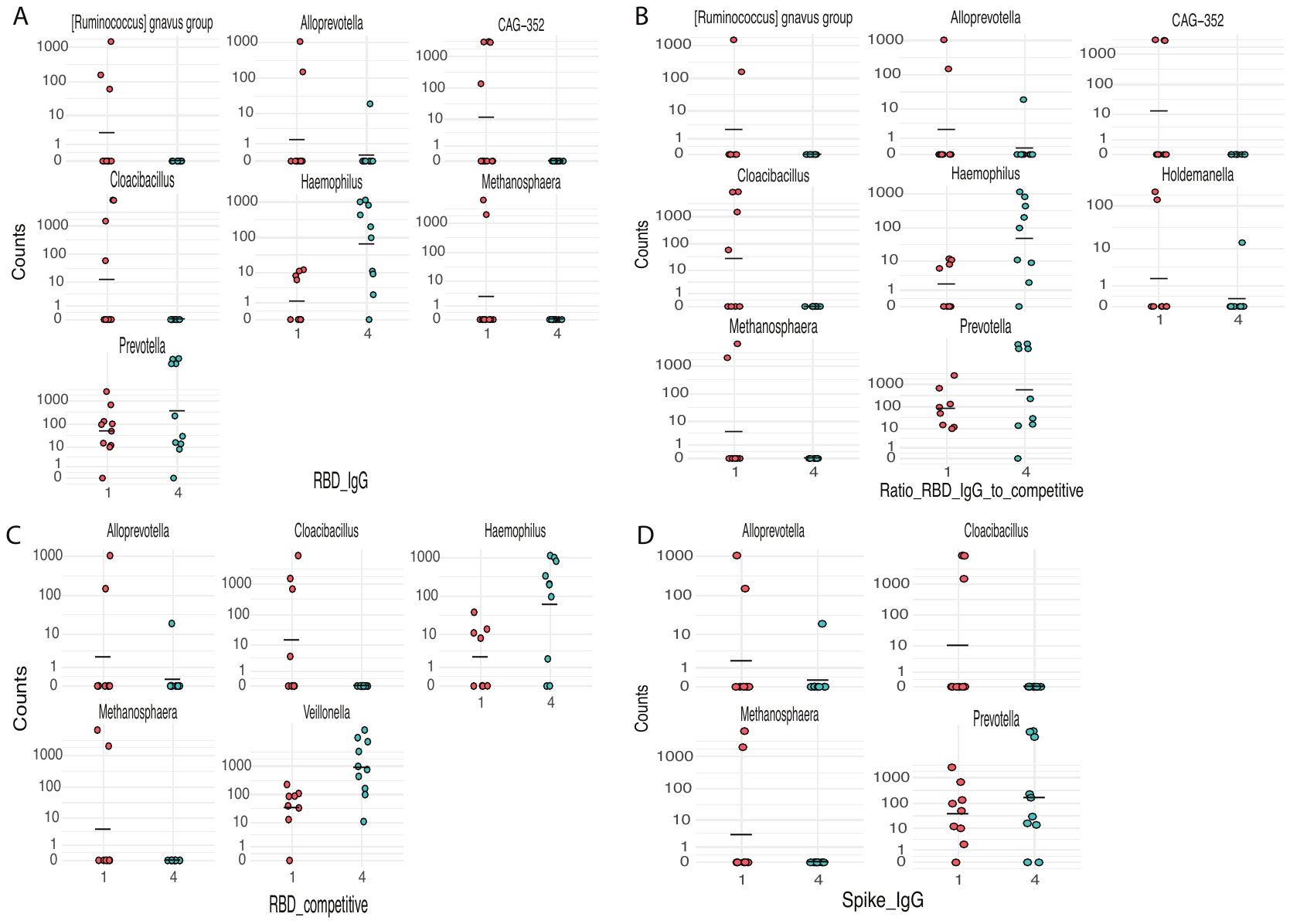

**Supplementary Figure 4** Significant differences in baseline microbial taxa in participants with differential (lowest [quartile 1] versus highest [quartile 4]) immune parameter responses, twelve-weeks post 1^st^ BNT162b2 vaccine. (A) The specific gut microbiota taxa that were significantly different (p<0.05; Wald test) between participants with the lowest and highest RBD IgG levels twelve-weeks post 1^st^ BNT162b2 vaccine. (B) The specific gut microbiota taxa that were significantly different (p<0.05; Wald test) between participants with the lowest and highest ratio of RBD IgG to competitive binding antibody levels twelve-weeks post 1^st^ BNT162b2 vaccine. (C) The specific gut microbiota taxa that were significantly different (p<0.05; Wald test) between participants with the lowest and highest RBD competitive binding antibody levels twelve-weeks post 1^st^ BNT162b2 vaccine. (D) The specific gut microbiota taxa that were significantly different (p<0.05; Wald test) between participants with the lowest and highest spike IgG levels twelve-weeks post 1^st^ BNT162b2 vaccine.

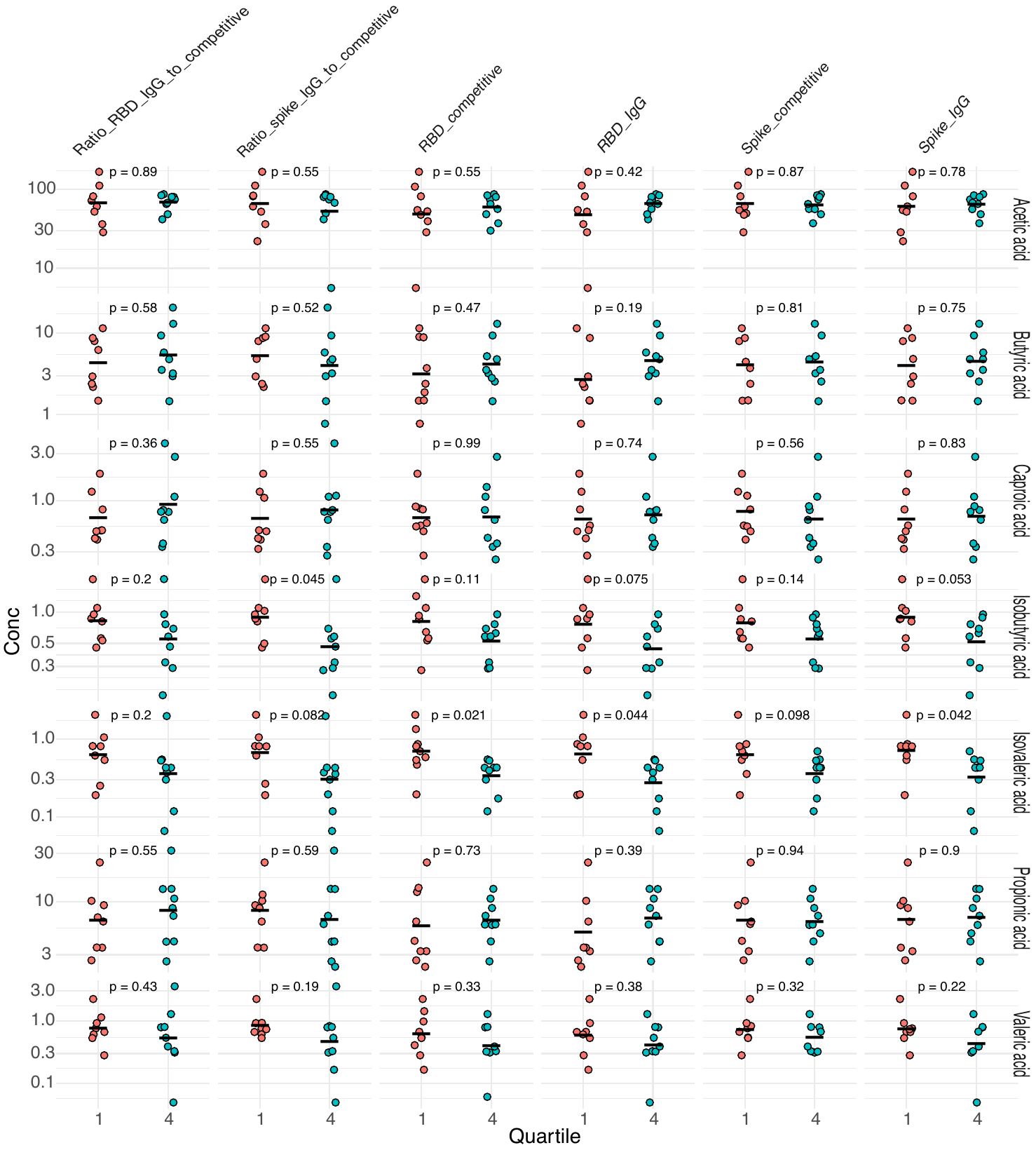

**Supplementary Figure 5** Significant (p<0.05; Wilcoxon) and non-significant differences in baseline SCFA concentrations (μmol/g stool) in participants with differential (lowest [quartile 1] versus highest [quartile 4]) immune parameter responses twelve-weeks post 1^st^ BNT162b2 vaccine.

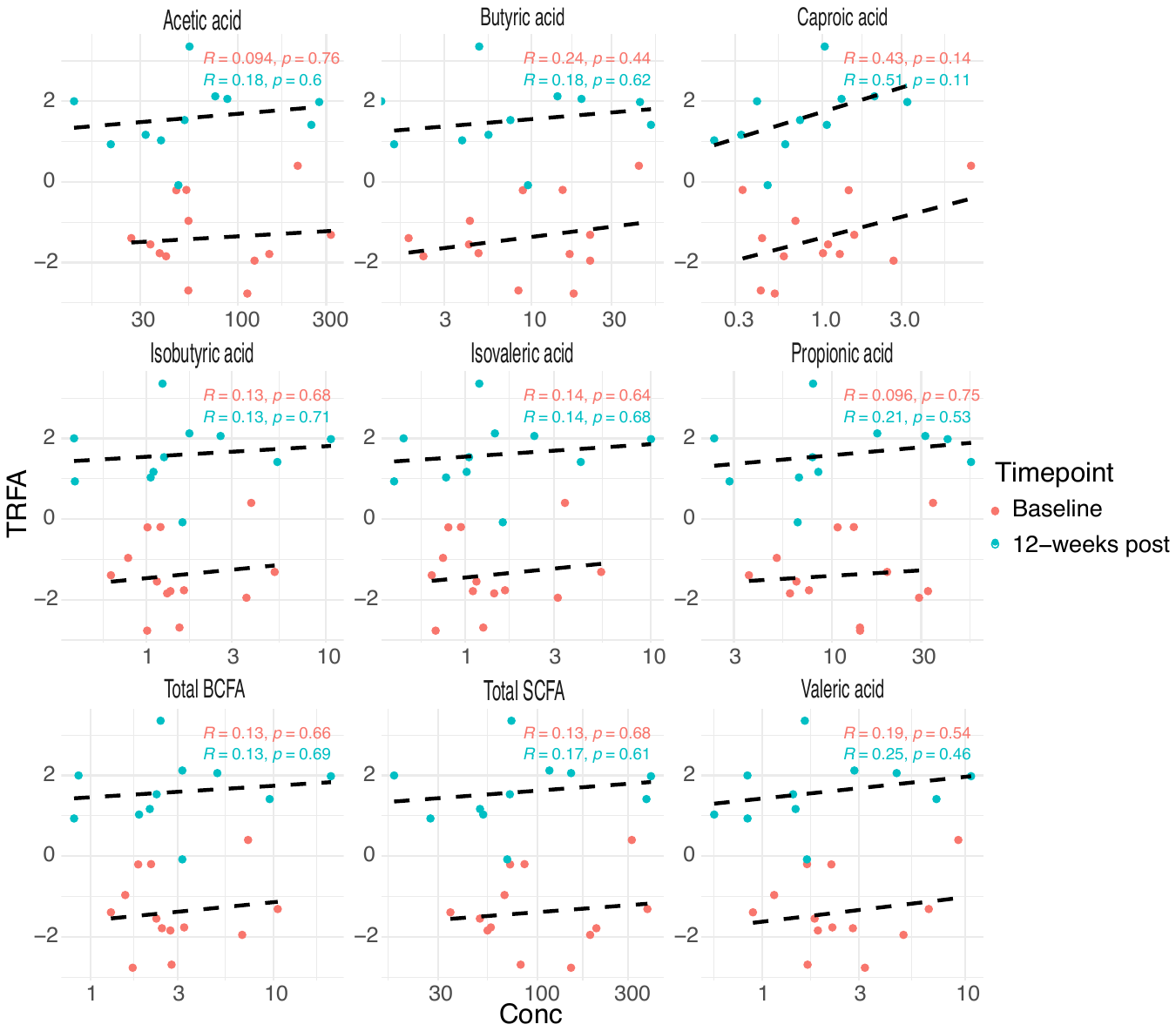

**Supplementary Figure 6** No significant correlation between total relative fractional avidity (TRFA) and baseline or twelve weeks post 1^st^ BNT162b2 vaccine SCFA concentrations (p>0.05; Spearman rank).

**Supplementary tables – refer to separate excel spreadsheet**
